# Sleep-related symptoms are associated with survival in oropharyngeal cancer radiotherapy survivors: Temporal characterization of patient-reported outcomes in a large-scale prospective longitudinal cohort

**DOI:** 10.1101/2025.09.20.25336187

**Authors:** Glennon Carevic, Rishabh Gaur, Warren Floyd, Cem Dede, Lucas McCullum, Cesar Marquez, Saadia A. Faiz, Mai Mandor Badawy, Amit Jethanandani, Elisabeta Marai, Abdallah Sherif Radwan Mohamed, Jacob Mathew, Guadalupe Canahuate, Naser Mohamed, Jack Phan, Sigmund Haidacher, Sarah Mirbahaeddin, Michael Spiotto, Anna Lee, Katherine Hutcheson, Amy Moreno, Clifton D. Fuller, MD Anderson Head and Neck Cancer Symptom Working Group

**Affiliations:** McGovern Medical School at University of Texas Houston Health Science Center, Houston, TX, USA; Department of Radiation Oncology, The University of Texas MD Anderson Cancer Center, Houston, TX, USA; University of Missouri-Kansas City School of Medicine, Kansas City, MO, USA; Department of Pulmonary Medicine, The University of Texas MD Anderson Cancer Center, Houston, TX, US; Department of Radiation Oncology, Winship Cancer Institute of Emory University, Atlanta, GA, USA; Department of Radiation Oncology, Baylor College of Medicine, Houston, TX, US; Department of Computer Science, University of Illinois Chicago, Chicago, IL, US; University of Houston, Houston, TX, USA; Quantitative Science Program, University of Texas MD Anderson Cancer Center UTHealth Houston Graduate School of Biomedical Sciences, Houston, TX, USA

**Keywords:** survival, oropharyngeal cancer, radiotherapy, sleep disturbance, fatigue, drowsiness

## Abstract

**Purpose:** Sleep-related symptom prevalence is not well-characterized and likely under-recognized in head and neck cancer. We investigated longitudinal symptom severity trajectory, prevalence of moderate/severe symptoms, and association between severe sleep-related patient-reported symptoms and oncologic endpoints in oropharyngeal cancer (OPC) patients receiving radiotherapy (RT).

**Methods:** A longitudinal cohort of 372 patients with OPC treated with curative-intent RT with/without chemotherapy at a single-site tertiary center was queried at RT start, end of RT, and six months post-RT. All completed baseline MD Anderson Symptom Inventory-Head and Neck (MDASI-HN) at start of RT. Sleep-related symptom items included “fatigue”, “disturbed sleep”, and “drowsiness” (mild, moderate, severe). Clinical data were collected, and overall survival (OS) was evaluated using Kaplan-Meier/log-rank tests and multivariable Cox proportional hazards models with covariate adjustment.

**Results:** Severe fatigue, disturbed sleep, and drowsiness were infrequent at baseline and at RT start, but peaked at RT end (26%, 15% and 18%) and returned to near-baseline prevalence by six months after RT (6%, 4%, 2%). In Cox proportional hazard models, severe longitudinal fatigue (HR 5.05, 95% CI 1.25-17.07, p = 0.013), sleep (HR 3.86, 95% CI 1.23 to 10.16, p = 0.0104), and drowsiness burden (HR 8.24, 95% CI 1.22 to 32.62, p = 0.008) were all independently associated with worse OS. Additionally, severe fatigue (HR = 3.58, 95% CI: 1.09 to 10.11, p = 0.0375), sleep disturbance (HR 5.08, 95% CI 1.60-13.51, p = 0.0081), and drowsiness (HR=7.76, 95% CI 1.45-33.47, p=0.0193) reported at RT start were also independently associated with significantly worse OS. Aside from age, severe average daily symptom scores, and severe symptoms reported at RT start, were independently associated with worse OS.

**Conclusion:** Severe sleep related symptoms were uncommon at baseline or RT start, but a substantial proportion of OPC patients experienced clinically meaningful sleep disturbances by the end of RT. At an average follow up of 84 months, severe longitudinal fatigue, sleep, and drowsiness burden were all independently associated with worse OS in OPC. Severe fatigue, drowsiness and sleep disturbance reported at RT start were also independently associated with worse OS. Assessment of sleep disturbances along with potential physiologic correlates are an important part of comprehensive care.

## 1 Introduction

Head and neck squamous cell carcinoma (HNSCC) is the seventh most common cancer worldwide, and it accounts for approximately 4.5% of all cancer diagnoses and 4.6% of all cancer-related mortalities.^1^ Per the American Cancer Society’s annual report, there will be over 58,000 new cases and over 12,000 deaths from oral cavity and pharyngeal cancers in the United States in 2024 (Cancer Facts and Figures). Over 90% of oropharyngeal cancers (OPC) are squamous cell carcinomas.^2–4^ A significant proportion of OPC is associated with human papillomavirus (HPV), and the overall prognosis is favorable with management by radiation therapy (RT) with or without chemotherapy.^5^ While improvements in RT techniques such as intensity-modulated radiotherapy (IMRT) have demonstrated a marked reduction in collateral damage to surrounding normal organs and tissues, areas outside the region-of-interest (ROI) still incur damages that could translate to detrimental effects on quality of life (QOL).^6^

Radiation toxicity to healthy body tissue can lead to a variety of side effects, including acute phase mucosal and orodental effects.^7,8^ Sleep disturbances are highly prevalent among patients with head and neck cancer, and these are associated with worse quality of life, increased fatigue and more severe treatment-related toxicities.9 These are notable during both the acute treatment phase and the “late” post-radiation period, with incidence as high as 75% of cases during and after RT.^10–12^ Interestingly, in a prospective, multisite cohort (n=457), Shuman and colleagues showed that sleep scores did not change dramatically from the time of diagnosis to 1 year after treatment, thus suggesting that those with poor sleep at baseline may be more susceptible to worsened sleep with treatment.^11^

Uninterrupted sleep has vital functions such as development, energy conservation, brain waste clearance, modulation of immune responses, cognition, performance, disease vigilance, and psychological state.^13,14^ Short-term sleep disturbances may cause increased stress responsivity, somatic pain, reduced quality of life, emotional distress, mood disorders, and deficits in cognition, memory, and performance deficits.14,15 Long-term effects of sleep disturbances on health include hypertension, dyslipidemia, cardiovascular disease, metabolic syndrome, type 2 diabetes, and many malignancies including breast, prostate, and colorectal cancer.^14,15^

Fatigue, drowsiness, and sleep disturbances are well-documented toxicities of RT for OPC.^6,16^ Among these, fatigue is the most extensively studied, though unlike drowsiness, it is not relieved by resting or sleeping.^17^ When left unaddressed, these symptoms can significantly disrupt treatment, potentially necessitating changes in RT dosing and fractionation, treatment interruption, or treatment discontinuation altogether.^6^ Identifying patients at risk of RT-induced sleep disturbances is challenging, given their multifactorial etiology.^18,19^

Prospective data on sleep disruption in patients with OPC are limited, and the mechanisms underlying these disturbances remain poorly understood. To address this, we have sought to ascertain, in a longitudinal prospective cohort, the relative prevalence, severity over time, and mortality impact of sleep-related patient-reported symptoms. OPC patients at our facility complete the MD Anderson Symptom Index (MDASI) as a component of their baseline assessment and for ongoing survivorship monitoring.

The MDASI head and neck (MDASI-HN) questionnaire subscale, a validated PRO measurement, quantifies the severity of head and neck cancer-related symptoms and their interference with daily activities. Through analysis of this data, our study aims to: (1) identify and assess the impact of the correlates of these symptoms, including demographics, disease characteristics, and treatment-related variables on survival outcomes; (2) track longitudinal changes in symptomatology; and (3) assess the impact of timepoint-specific and overall symptom burden in order to identify OPC patient subgroups who may benefit from early supportive care interventions.

## 2 Methods

### 2.1 Study Design

A retrospective cohort study of prospectively collected data from patients with OPC treated at the University of Texas MD Anderson Cancer Center (MDACC) was conducted. This study was performed under an Institutional Review Board-approved protocol, with data derived from a prospectively maintained symptom outcomes registry for patients with head and neck cancer. We adhered to STROBE (Strengthening the Reporting of Observational Studies in Epidemiology) guidelines in study design and reporting.

Records of patients enrolled in the MDACC Oropharynx Cancer Program database, a program generously supported by Mr. and Mrs. Charles W. Stiefel (Institutional Protocol PA14-0947), were reviewed. This database includes all consecutive patients with OPC and squamous cell carcinoma in the neck (unknown primaries) evaluated and/or treated at MDACC who were willing to provide consent to be included in the database since March 2015. This robust database tracks patient demographics such as age, gender, smoking history, HPV-status, and cancer stage. Chemotherapy regimen(s) and radiation treatment dosing and technique as well as response to therapy and survival are tracked. Patient outcomes are also prospectively tracked by the Patient-Reported Outcomes/Function (PROF) core as well as toxicities using the MDASI-HN.

### 2.2 Patient Cohort

Pathologically confirmed patients with OPC who completed a curative intent course of RT at MDACC were eligible for the study. All included patients completed the MDASI-HN questionnaire at the start of RT. Patients were excluded if they: (1) had a previous history of RT to the head and neck or (2) did not have MDASI-HN data at the start of RT. Data were continuously prospectively collected and stored in an existing electronic database.

### 2.3 MDASI-HN Questionnaire Subscale

Patient-reported outcomes (PROs) were assessed using the MDASI-HN questionnaire subscale, a validated 28-item tool for measuring patient-reported symptoms and their impact. This specialized tool comprises three distinct components: 13 core items that assess the severity of symptoms commonly across all cancer types, 6 items that evaluate the degree to which symptoms interfere with a patient’s ability to perform daily activities, and 9 items specifically chosen to rate symptoms unique to head and neck cancer. The 13 core and 9 HN specific items are both rated on a 0-10 numeric scale, with 0 representing “not present” and 10 signifying “as bad as you can imagine,” based on the worst severity experienced. Interference items are rated on a 0-10 scale ranging from “did not interfere” to “interfered completely,” indicating the extent to which symptoms disrupt the patient’s ability to carry out daily activities. The combination of these three components allows the MDASI-HN to provide a comprehensive evaluation of symptom burden in patients with head and neck cancer. The MDASI-HN questionnaire data are a subset of the MDACC Oropharynx Cancer Program database, which provides an opportunity to analyze RT-induced fatigue, drowsiness, and sleep disturbances in this cohort.

### 2.4 Statistical Analysis

Descriptive statistics were calculated for patients’ (1) demographics (2) tumor characteristics (3) treatment characteristics and (4) MDASI-HN item scores. A separate multiple linear regression model using standard least squares was fit for each symptom to assess the association between cumulative symptom burden and clinical covariates. Predictor variables included age, sex, tumor site, tumor stage, nodal status, induction chemotherapy, and surgery. MDASI-HN individual items (fatigue; sleep disturbance; drowsiness) were coded using a 10-point scale where a score of 0 corresponds to “not present” and a score of 10 corresponds to “as bad as you can imagine.” MDASI-HN questionnaires for symptom assessment item scores were converted into mild (score: 0-3), moderate (score: 4-6), and severe (score: 7-10) categories.

To quantify cumulative symptom burden over time, we calculated patient-level AUC values for fatigue, sleep disturbance, and drowsiness using the trapezoidal integration rule. For each patient, raw AUC values were averaged by dividing by the interval from RT start to the last available MDASI-HN survey, yielding an average daily symptom score. Average daily symptom scores were categorized using thresholds adapted from standard MDASI severity cutoffs: mild (0-3), moderate (4-6), and severe (7-10).

For survival analyses, we performed univariate comparisons of OS using Kaplan-Meier survival curves, stratified by symptom severity categories (mild, moderate, severe) at key timepoints. The log-rank test was used for pairwise comparisons between severity groups. We then constructed multivariate Cox proportional hazards models to evaluate whether symptom severity was independently associated with OS, adjusting for potential confounders.

Multivariate Cox proportional hazards models were fit for each symptom domain (fatigue, sleep disturbance, drowsiness) at the start of RT. Covariates included age (continuous), sex, T stage (categorical T0-T4), nodal status (N0 vs. N+), induction chemotherapy (yes/no), and surgery prior to RT (yes/no). Additional multivariate Cox models were fit using the binned average daily symptom scores for each symptom domain (fatigue, sleep disturbance, drowsiness). The same covariates from the previous Cox models were included. The proportional hazards (PH) assumption was tested for all Cox models (based on Schoenfeld residuals and time interactions); no violations were observed. Hazard ratios (HRs) with 95% confidence intervals (CIs) and two-sided p-values were reported, with p < 0.05 considered statistically significant.

We calculated each patient’s symptom burden as a percentage of the maximum possible AUC (i.e. area under the curve across all timepoints divided by the area corresponding to a constant score of 10 at all timepoints). Using these patient-level burden values, we generated heat maps of symptom burden stratified by RT-start severity, cumulative distribution curves of patients exceeding given burden thresholds, and follow-up cutoff analyses to see if group differences persisted over time. We then generated pairwise scatterplots with r2 values to analyze the relationship between fatigue, sleep, and drowsiness.

Statistical analyses were performed using the JMP Statistical Software Version 17.1 (Cary, NC), GraphPad Prism 10.3.1 and lifelines in Python.

## 3 Results

### 3.1 Patients

A total of 372 OPC patients who received a curative course of RT at MDACC completed MDASI-HN questionnaires at the start of RT and other time points. Table 1 summarizes the cohort demographics and clinical characteristics. The median age of participants was 60 years (IQR 53-67), and 338 patients (91%) were male. Most primary tumors arose in the base of tongue (BOT) (49%) or tonsil (39%), with 12% at other oropharyngeal subsites. Concurrent chemoradiotherapy was administered in 285 patients (77%) and 52 patients (14%) received induction chemotherapy; 20% of patients had undergone surgical resection of the primary tumor before RT. Most patients had low-volume primary tumors (e.g. T1 or T2; 67%), 37% had clinically N1 disease per the American Joint Commission on Cancer (AJCC) 7th edition staging criteria. All patients received RT to a median treatment prescription of 69.96 Gy in 33 fractions, via IMRT/IMPT.

**Table 1.**
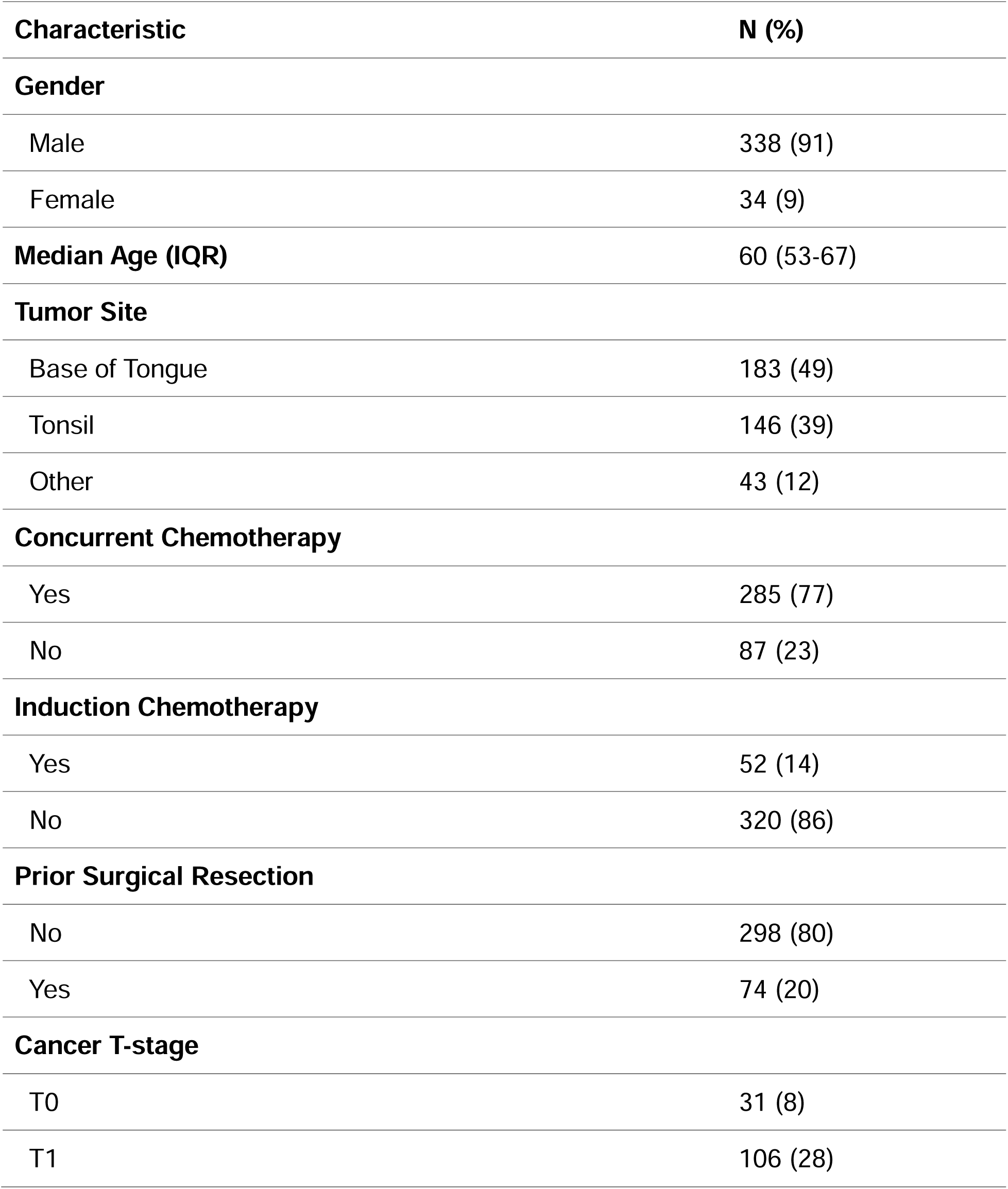
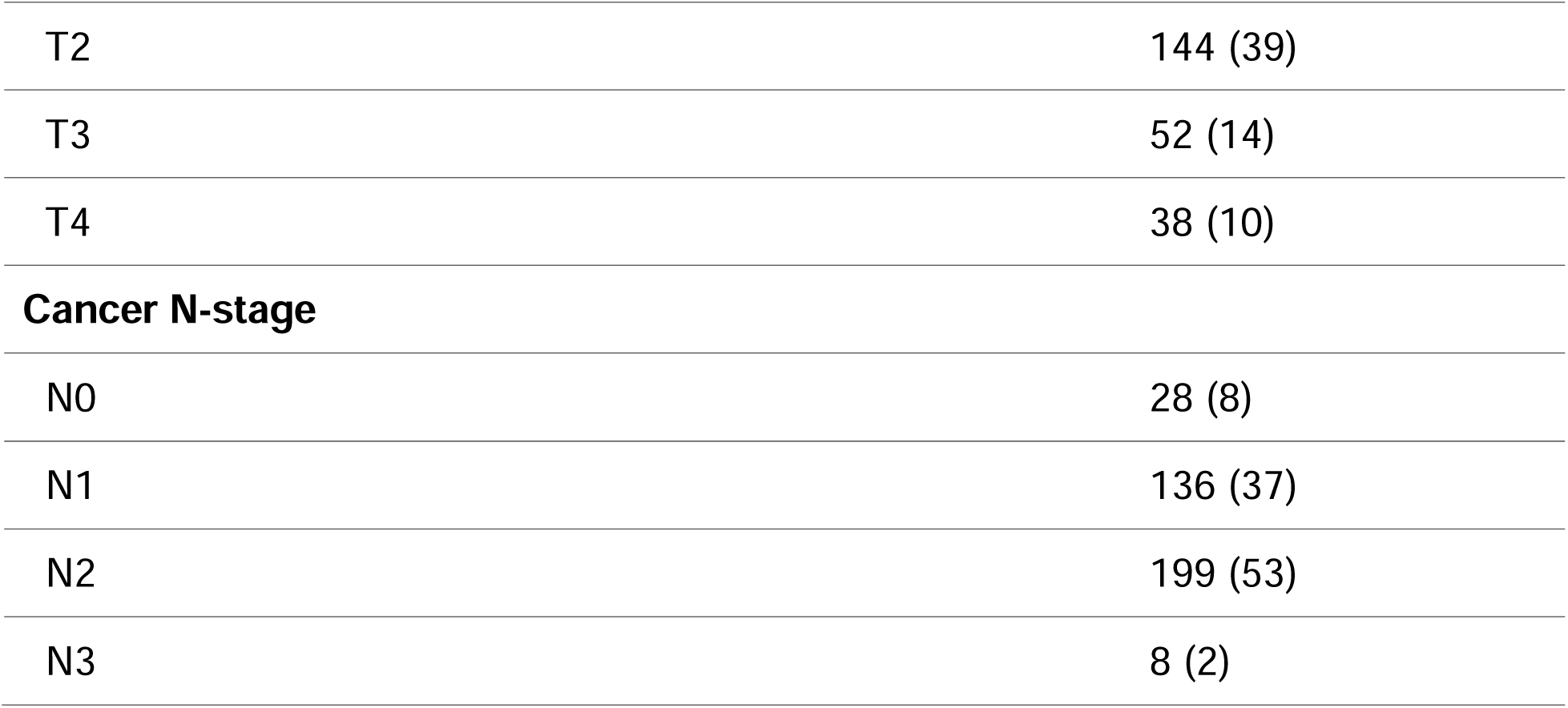
Patient Demographics.

| Characteristic | N (%) |
| --- | --- |
| **Gender** |  |
| Male | 338 (91) |
| Female | 34 (9) |
| **Median Age (IQR)** | 60 (53-67) |
| **Tumor Site** |  |
| Base of Tongue | 183 (49) |
| Tonsil | 146 (39) |
| Other | 43 (12) |
| **Concurrent Chemotherapy** |  |
| Yes | 285 (77) |
| No | 87 (23) |
| **Induction Chemotherapy** |  |
| Yes | 52 (14) |
| No | 320 (86) |
| **Prior Surgical Resection** |  |
| No | 298 (80) |
| Yes | 74 (20) |
| **Cancer T-stage** |  |
| T0 | 31 (8) |
| T1 | 106 (28) |
| T2 | 144 (39) |
| T3 | 52 (14) |
| T4 | 38 (10) |
| **Cancer N-stage** |  |
| N0 | 28 (8) |
| N1 | 136 (37) |
| N2 | 199 (53) |
| N3 | 8 (2) |

**Table 1.**
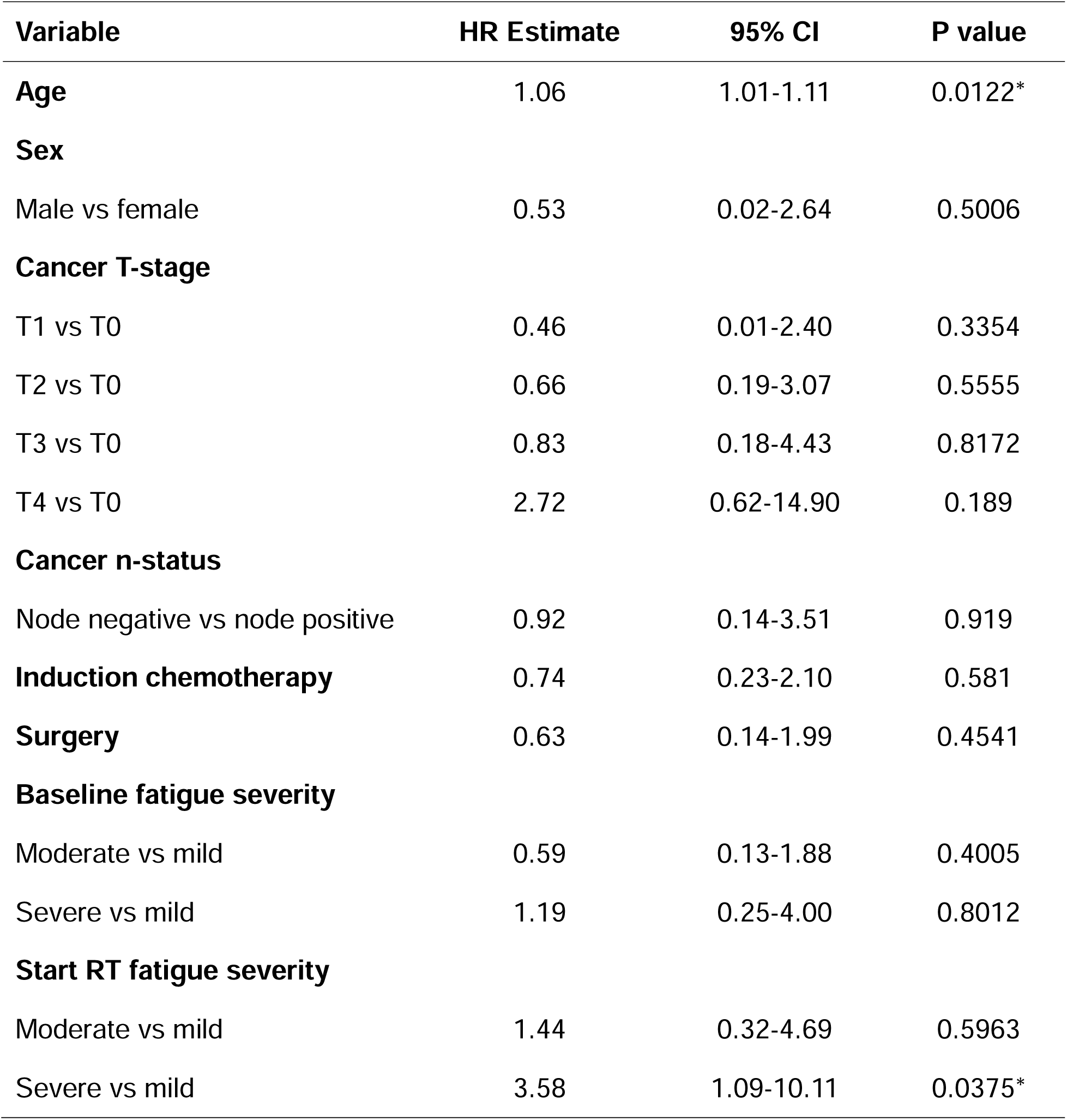
Cox proportional hazards analysis for fatigue at specific timepoints.

Patient compliance with PRO assessments was high at RT initiation and remained high at subsequent timepoints: 84% completed the MDASI-HN questionnaire at baseline, 79% at end of RT, and 77% at 6 months post-RT, with diminishing response rates at later follow-ups as expected in a long-term cohort.

In the multiple linear regression model using standard least squares for average daily fatigue scores, T0 stage (coefficient: 1.58, 95% CI: 0.37 to 2.78, p = 0.011; S1) and primary tumors located in BOT (coefficient: 0.78, 95% CI: 0.07 to 1.49, p = 0.032) were significantly associated with fatigue burden. All other predictors, including age, sex, nodal status, induction chemotherapy, and surgery were not significantly associated with fatigue. In the model for average daily sleep scores, T0 stage was significantly associated with increased sleep burden (coefficient: 1.11, 95% CI: 0.01 to 2.21, p = 0.047; S2), while all other variables were not significant. For average daily drowsiness scores, prior surgical resection was significantly associated with drowsiness burden (coefficient = 0.21, 95% CI: 0.00 to 0.42, p = 0.049; S3), whereas age, sex, tumor site, tumor stage, nodal status, and induction chemotherapy were not significant. We calculated combined average daily symptom scores for fatigue, sleep, and drowsiness. In the model, only T0 stage was significantly associated with combined symptom burden (coefficient = 1.05, 95% CI: 0.07 to 2.03, p = 0.037; S4).

### 3.2 Fatigue

At baseline, fatigue was mostly mild (82% of patients; 12% moderate, 6% severe; mean score 1.8). This distribution was unchanged at RT start (mean 1.9; S5, S6). At the start of RT, fatigue severity was largely unchanged from baseline: 82% reported mild fatigue, 12% moderate, and 6% severe (mean score 1.9). Fatigue worsened during RT, peaking by the end of treatment: only 39% reported mild fatigue, while 36% reported moderate, and 26% reported severe fatigue. The mean fatigue score increased to 4.5 at end of RT. Following treatment, fatigue gradually improved. By 6 months post-RT, the distribution of fatigue severity had largely returned to baseline levels (approximately 83% mild, 12% moderate, 6% severe, mean score ∼1.9), and these proportions remained stable at 12 months post-RT and beyond.

The distribution of patient-reported fatigue symptoms over time, showing the percentage of patients with mild (blue), moderate (green), or severe (red) fatigue at each timepoint during and after RT are shown in Figure 1. Patients with mild fatigue reported at the start of treatment largely remained in the mild category across timepoints, while those with moderate or severe fatigue experienced greater symptom fluctuation, with peak worsening during the end of RT and partial recovery by 6-12 months.

**Figure 1.**
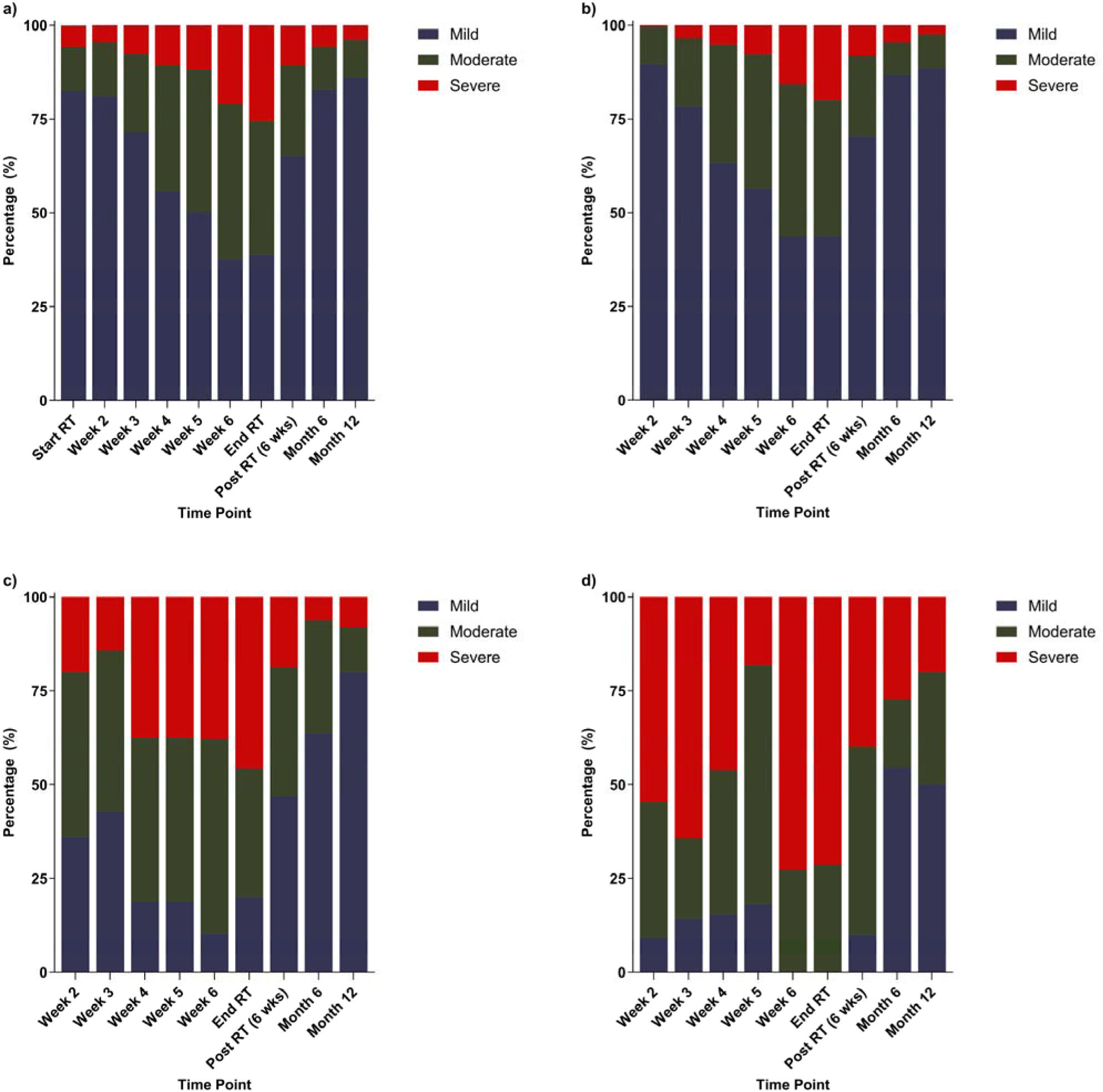
Evolution of fatigue severity at relevant timepoints, stratified by symptom severity at RT start. Panels represent: (a) the entire cohort, (b) patients with mild fatigue at RT start, (c) patients with moderate fatigue at RT start, and (d) patients with severe fatigue at RT start.

A multivariate Cox proportional hazards model was used to evaluate the independent association of patient-reported symptom severity with OS, adjusting for potential confounders. Patients who reported severe fatigue symptoms at the start of RT had significantly worse OS compared to those with mild symptoms (HR = 3.58, 95% CI: 1.09 to 10.11, p = 0.0375; Table 2). Moderate fatigue at RT start was not significantly associated with OS (HR = 1.44, 95% CI 0.32-4.69; p = 0.60). Aside from age, no other variable (including moderate fatigue at RT initiation or moderate/severe fatigue at the end of RT or other timepoints) was independently associated with OS in this model. A separate Cox model was used to evaluate the independent association of average daily fatigue scores, adjusting for the same potential confounders. Patients with severe longitudinal fatigue burden had significantly worse OS compared with those with mild burden (HR 5.05, 95% CI 1.25-17.07, p = 0.013), whereas moderate burden was not significantly associated with outcome (HR 0.77, 95% CI 0.25-1.96, p = 0.6101; Table 3). Aside from age, no other variable was independently associated with OS in this model.

**Table 2.** Cox proportional hazards analysis for average daily fatigue.

| Variable | HR Estimate | 95% CI | P value |
| --- | --- | --- | --- |
| **Age** | 1.07 | 1.02 to 1.12 | 0.0052* |
| **Sex** |  |  |  |
| Male vs female | 1.45 | 0.38-9.73 | 0.6385 |
| **Cancer T-stage** |  |  |  |
| T1 vs T0 | 0.42 | 0.09-2.15 | 0.2579 |
| T2 vs T0 | 0.76 | 0.23-3.43 | 0.6784 |
| T3 vs T0 | 0.69 | 0.15-3.56 | 0.6278 |
| T4 vs T0 | 1.49 | 0.28-8.96 | 0.6431 |
| **Cancer n-status** |  |  |  |
| Node negative vs node positive | 0.89 | 0.14-3.35 | 0.8777 |
| **Induction chemotherapy** | 0.49 | 0.12-1.67 | 0.2902 |
| **Surgery** | 0.58 | 0.13-1.76 | 0.4541 |
| **Concurrent chemotherapy** | 3.34 | 1.07-15.11 | 0.0659 |
| **Average daily fatigue severity** |  |  |  |
| Moderate vs mild | 0.77 | 0.25-1.96 | 0.6101 |
| Severe vs mild | 5.05 | 1.25-17.07 | 0.0130* |

**Table 3.** Multivariate Cox proportional hazards analysis for sleep.

| Variable | HR Estimate | 95% CI | P value |
| --- | --- | --- | --- |
| **Age** | 1.06 | 1.01-1.11 | 0.0127* |
| **Sex** |  |  |  |
| Male vs female | 0.31 | 0.01-1.66 | 0.2009 |
| **Cancer T-stage** |  |  |  |
| T1 vs T0 | 0.46 | 0.10-2.45 | 0.341 |
| T2 vs T0 | 0.59 | 0.17-2.75 | 0.4638 |
| T3 vs T0 | 0.84 | 0.18-4.45 | 0.8218 |
| T4 vs T0 | 1.81 | 0.430-9.64 | 0.4331 |
| **Cancer n-status** |  |  |  |
| Node negative vs node positive | 1.02 | 0.16-3.79 | 0.9838 |
| **Induction chemotherapy** | 0.88 | 0.27-2.53 | 0.8236 |
| **Surgery** | 0.57 | 0.13-1.75 | 0.3503 |
| **Baseline sleep severity** |  |  |  |
| Moderate vs mild | 0.62 | 0.03-3.38 | 0.6341 |
| Severe vs mild | 1.64 | 0.49-4.54 | 0.3991 |
| **Start RT sleep severity** |  |  |  |
| Moderate vs mild | 0.62 | 0.09-2.52 | 0.5382 |
| Severe vs mild | 5.08 | 1.60-13.51 | 0.0081* |

Kaplan-Meier analysis was performed to evaluate OS in fatigue severity categories, followed by pairwise log-rank tests to evaluate the difference in OS between mild vs moderate and moderate vs severe fatigue categories. Pairwise log-rank tests revealed that the difference in survival between the moderate and severe groups was significantly different (p = 0.0157). The difference between mild and moderate fatigue groups was not statistically significant (p = 0.8768). The Kaplan-Meier survival curves for all 3 groups are displayed in Figure 2. A separate Kaplan-Meier analysis was performed to evaluate OS in average daily fatigue scores, stratified by severity (Figure 3).

**Figure 2.**
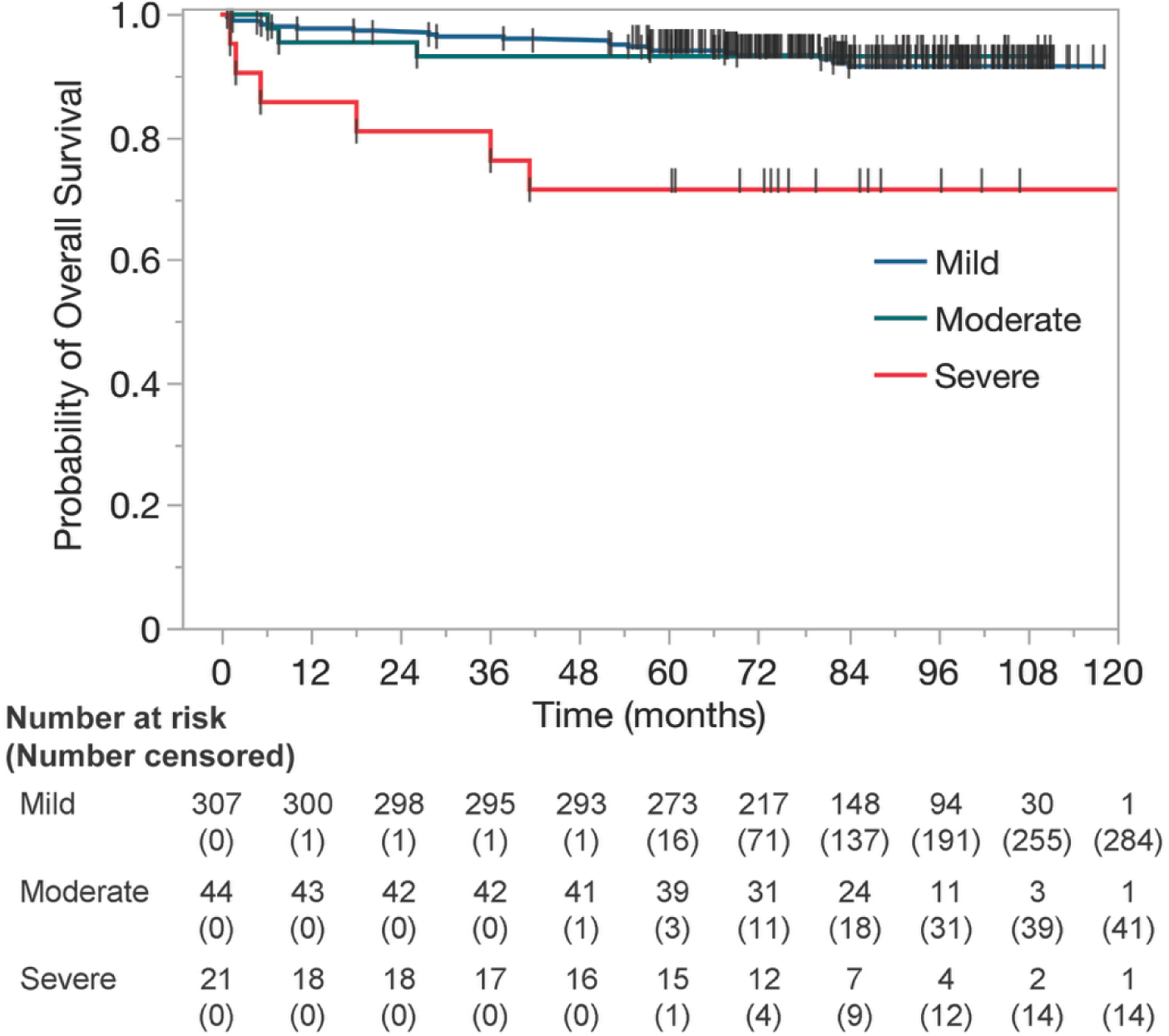
Kaplan-Meier survival curves stratified by fatigue severity at the start of RT

**Figure 3.**
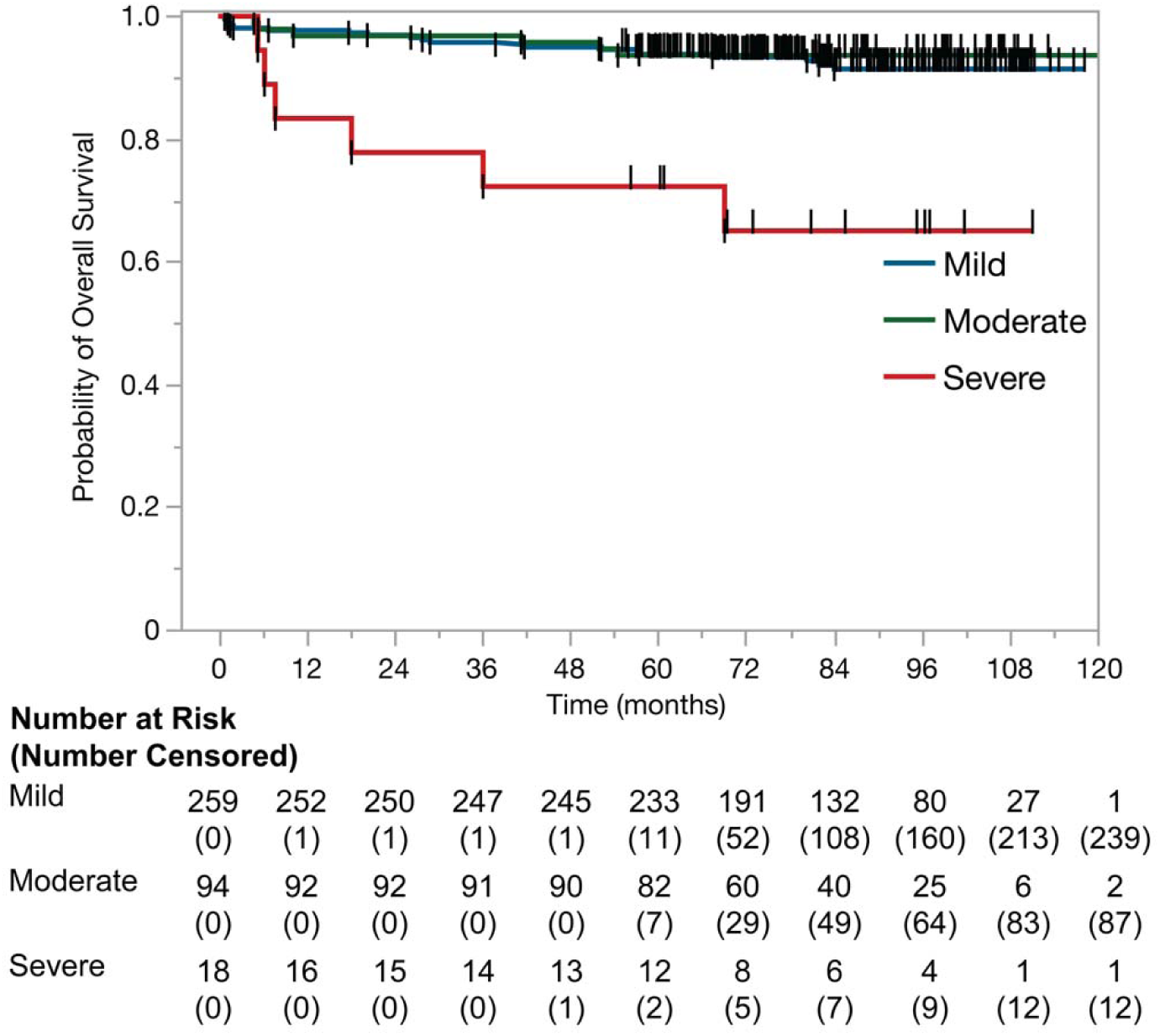
Kaplan-Meier survival curves stratified by severity of average daily fatigue scores

### 3.3 Sleep

At baseline, 83% of patients reported mild sleep disturbance, 11% reported moderate, and 6% reported severe (S7). Mean MDASI score at this time was 1.6 (SD: 2.32; S1). At the start of RT with a mean MDASI score of 1.7 (SD 2.11), 84% of patients continued to report mild sleep disturbance, while 11% had moderate and 5% had severe sleep problems. By the end of RT, sleep issues worsened (mean score 3.5). By 6 months post-RT, sleep disturbance had largely abated to near baseline levels, with 87% reporting mild, 10% moderate, and 4% severe (mean score 1.5, SD 1.95).

Patients reporting mild sleep disturbance at the start of treatment largely remained in the mild category across timepoints (Figure 4). Those reporting moderate sleep symptoms experienced greater symptom fluctuation, with peak worsening during the end of RT and partial recovery by 6-12 months. Patients reporting severe sleep disturbance at the start of RT reported gradually less severe symptoms over the trajectory of the treatment.

**Figure 4.**
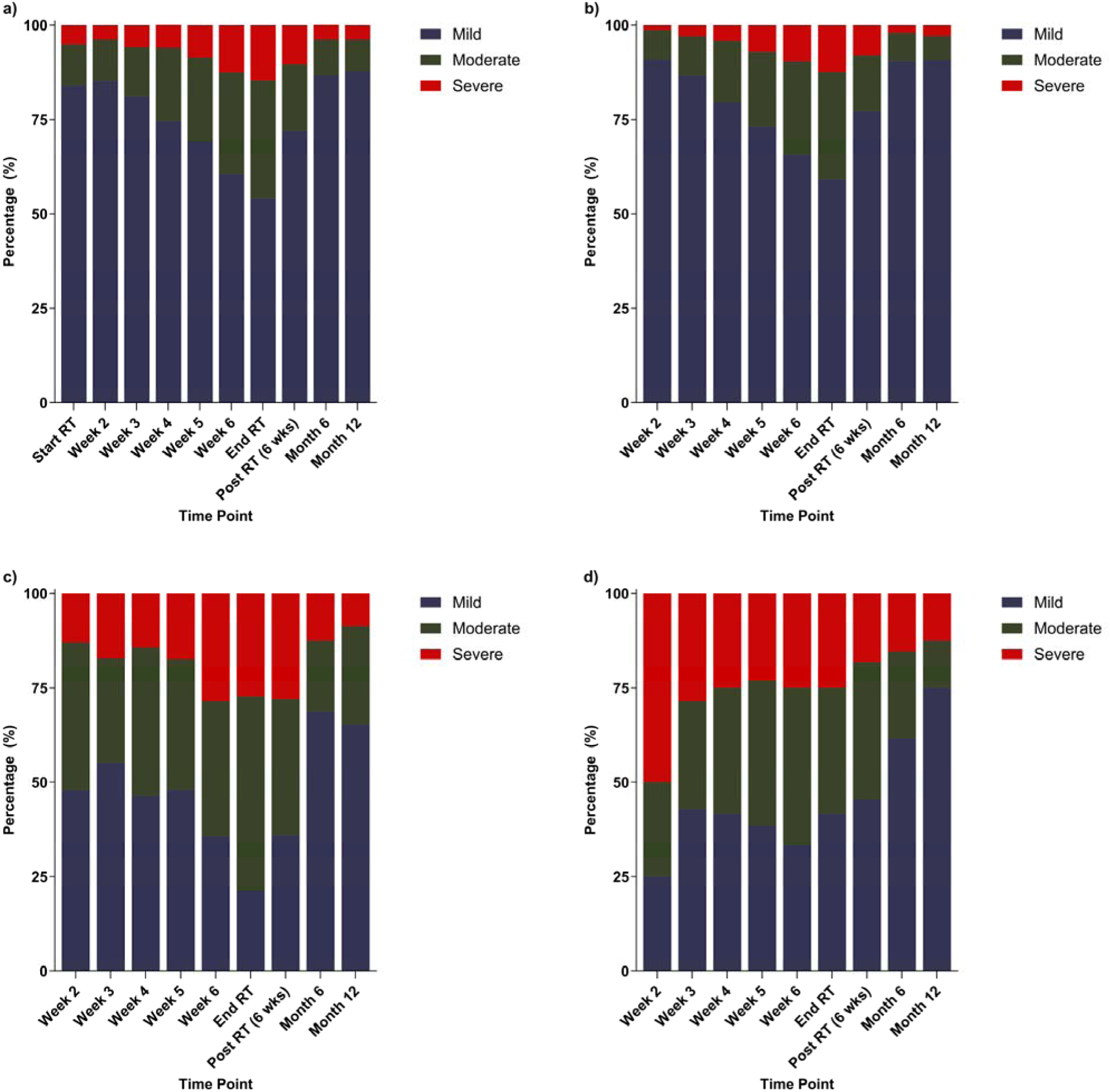
Evolution of sleep severity at relevant timepoints, stratified by symptom severity reported at RT start. Panels represent: (a) the entire cohort, (b) patients with mild sleep disturbance at RT start, (c) patients with moderate sleep disturbance at RT start

The results of the Cox proportional hazards models for sleep-related symptoms at baseline and the start of RT are shown in Table 4. At RT initiation, patients who reported severe levels of sleep disturbance had a fivefold higher risk of death compared to those with mild sleep symptoms (HR: 5.08 95% CI: 1.60 to 13.51, p = 0.0081). This association remained significant after controlling for age and other clinical factors, none of which were independently prognostic in the sleep disturbance model aside from age. A separate Cox model for average daily sleep scores showed that patients with severe longitudinal sleep burden had significantly poorer survival compared with those with mild burden (HR 3.86, 95% CI 1.23 to 10.16, p = 0.0104), whereas moderate burden was not significantly associated with outcome (HR 0.37, 95% CI 0.06 to 1.25, p = 0.1767; Table 5). Aside from age, no other variable was independently associated with OS.

**Table 4.**
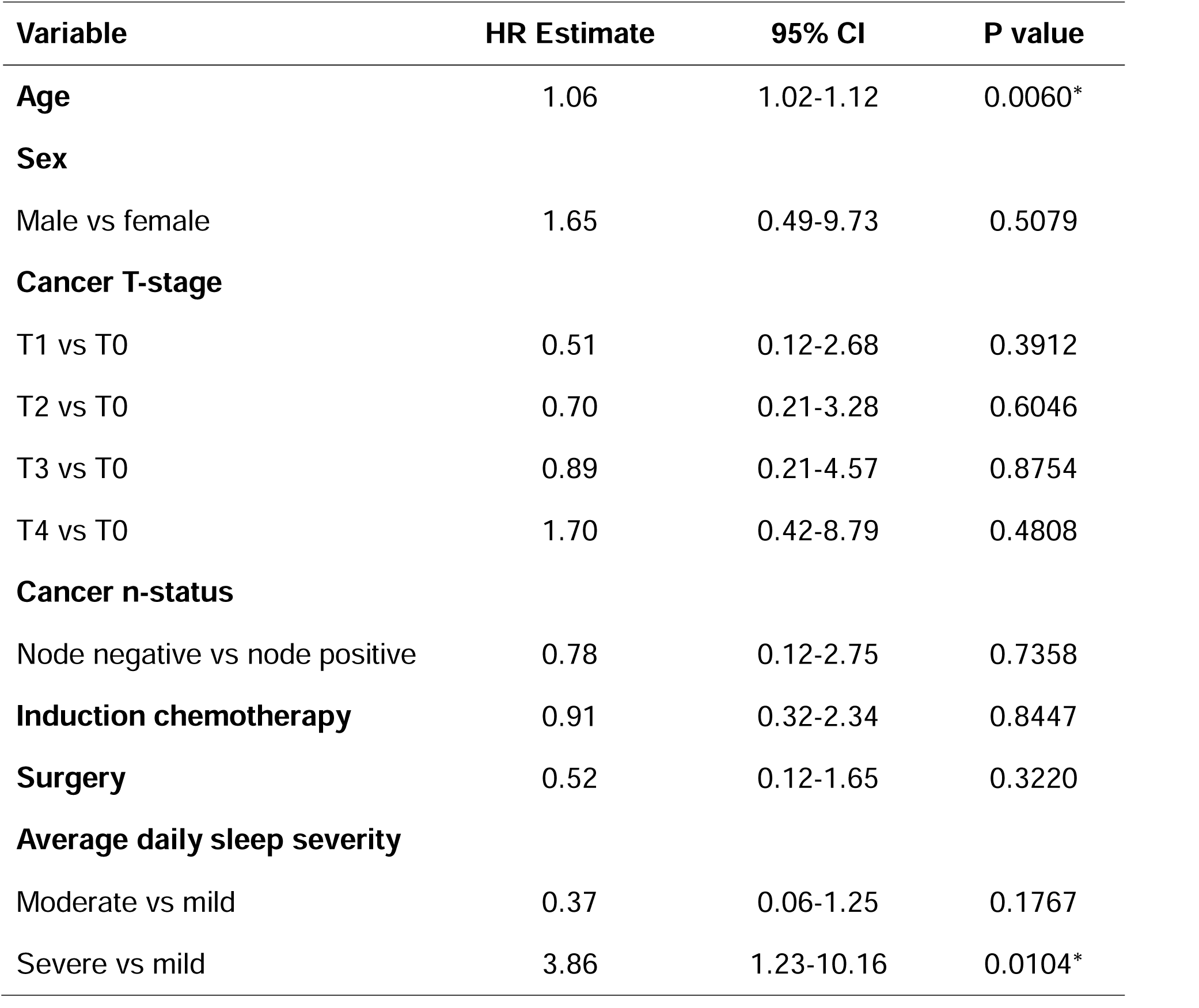
Multivariate Cox proportional hazards analysis for average daily sleep disturbance.

| Variable | HR Estimate | 95% CI | P value |
| --- | --- | --- | --- |
| **Age** | 1.06 | 1.02-1.12 | 0.0060* |
| **Sex** |  |  |  |
| Male vs female | 1.65 | 0.49-9.73 | 0.5079 |
| **Cancer T-stage** |  |  |  |
| T1 vs T0 | 0.51 | 0.12-2.68 | 0.3912 |
| T2 vs T0 | 0.70 | 0.21-3.28 | 0.6046 |
| T3 vs T0 | 0.89 | 0.21-4.57 | 0.8754 |
| T4 vs T0 | 1.70 | 0.42-8.79 | 0.4808 |
| **Cancer n-status** |  |  |  |
| Node negative vs node positive | 0.78 | 0.12-2.75 | 0.7358 |
| **Induction chemotherapy** | 0.91 | 0.32-2.34 | 0.8447 |
| **Surgery** | 0.52 | 0.12-1.65 | 0.3220 |
| **Average daily sleep severity** |  |  |  |
| Moderate vs mild | 0.37 | 0.06-1.25 | 0.1767 |
| Severe vs mild | 3.86 | 1.23-10.16 | 0.0104* |

**Table 5.** Multivariate Cox proportional hazards analysis for drowsiness.

| Variable | HR Estimate | 95% CI | P value |
| --- | --- | --- | --- |
| **Age** | 1.05 | 1.01-1.10 | 0.0138* |
| **Sex** |  |  |  |
| Male vs female | 0.29 | 0.02-1.64 | 0.1913 |
| **Cancer T-Stage** |  |  |  |
| T1 vs T0 | 0.34 | 0.07-1.87 | 0.2021 |
| T2 vs T0 | 0.65 | 0.18-3.06 | 0.5443 |
| T3 vs T0 | 0.85 | 0.18-4.51 | 0.8331 |
| T4 vs T0 | 1.92 | 0.46-10.3 | 0.3832 |
| **Cancer n-status** |  |  |  |
| Node negative vs node positive | 1.00 | 0.15-3.71 | 0.9980 |
| **Induction chemotherapy** | 0.96 | 0.29-2.76 | 0.9366 |
| **Surgery** | 0.55 | 0.12-1.70 | 0.3200 |
| **Baseline drowsiness severity** |  |  |  |
| Moderate vs mild | 0.30 | 0.02-1.78 | 0.2158 |
| Severe vs mild | 1.08 | 0.23-3.74 | 0.9089 |
| **Start RT drowsiness severity** |  |  |  |
| Moderate vs mild | 1.67 | 0.38-5.41 | 0.4467 |
| Severe vs mild | 7.76 | 1.45-33.47 | 0.0193* |

Kaplan-Meier analysis/pairwise log-rank tests revealed a statistically significant difference in survival between the moderate and severe groups (p = 0.0451), while the difference between mild and moderate drowsiness groups was not statistically significant (p = 0.9068) (Figure 5).

**Figure 5.**
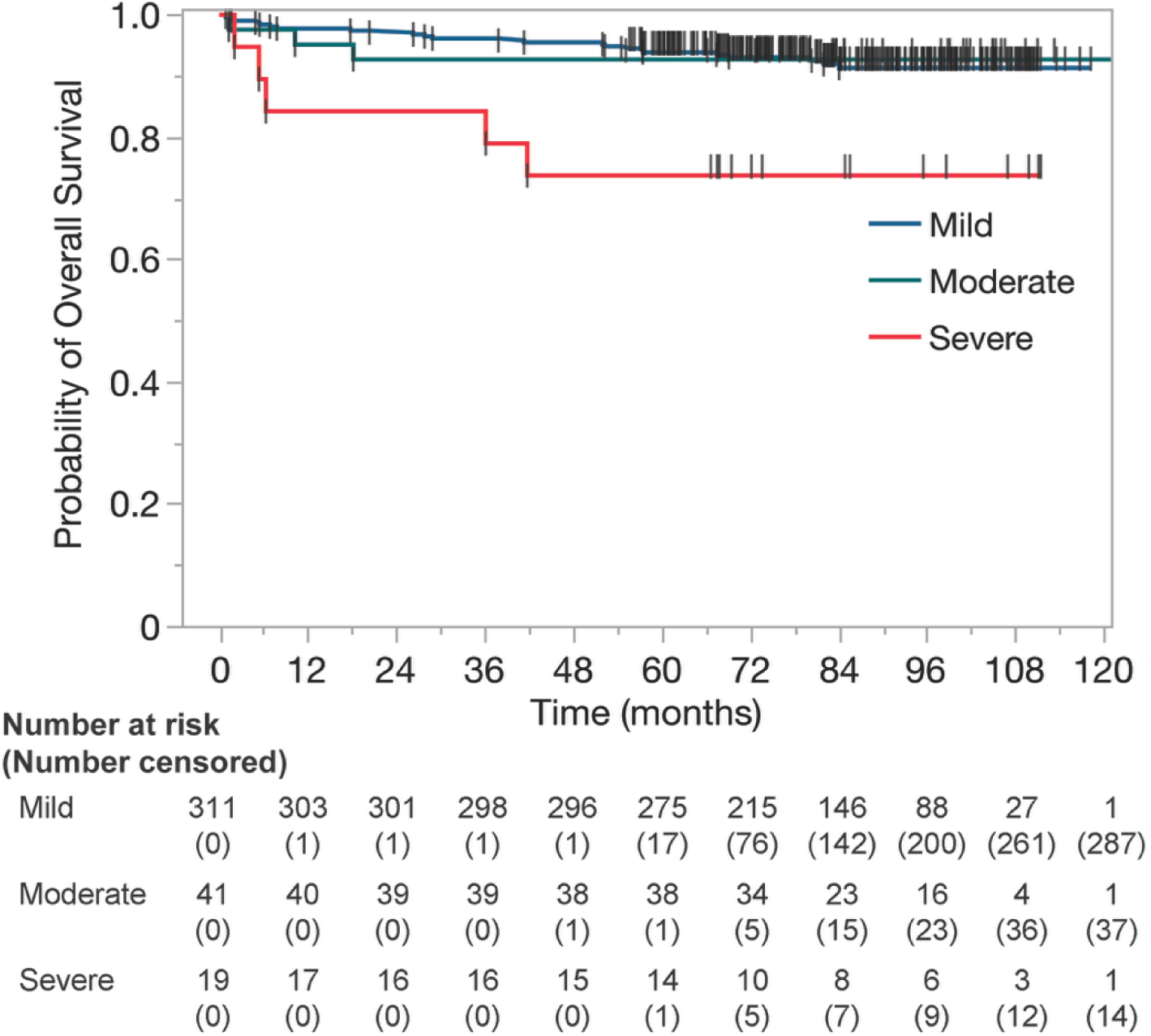
Kaplan-Meier survival curves stratified by sleep severity at the start of RT

**Figure 6.**
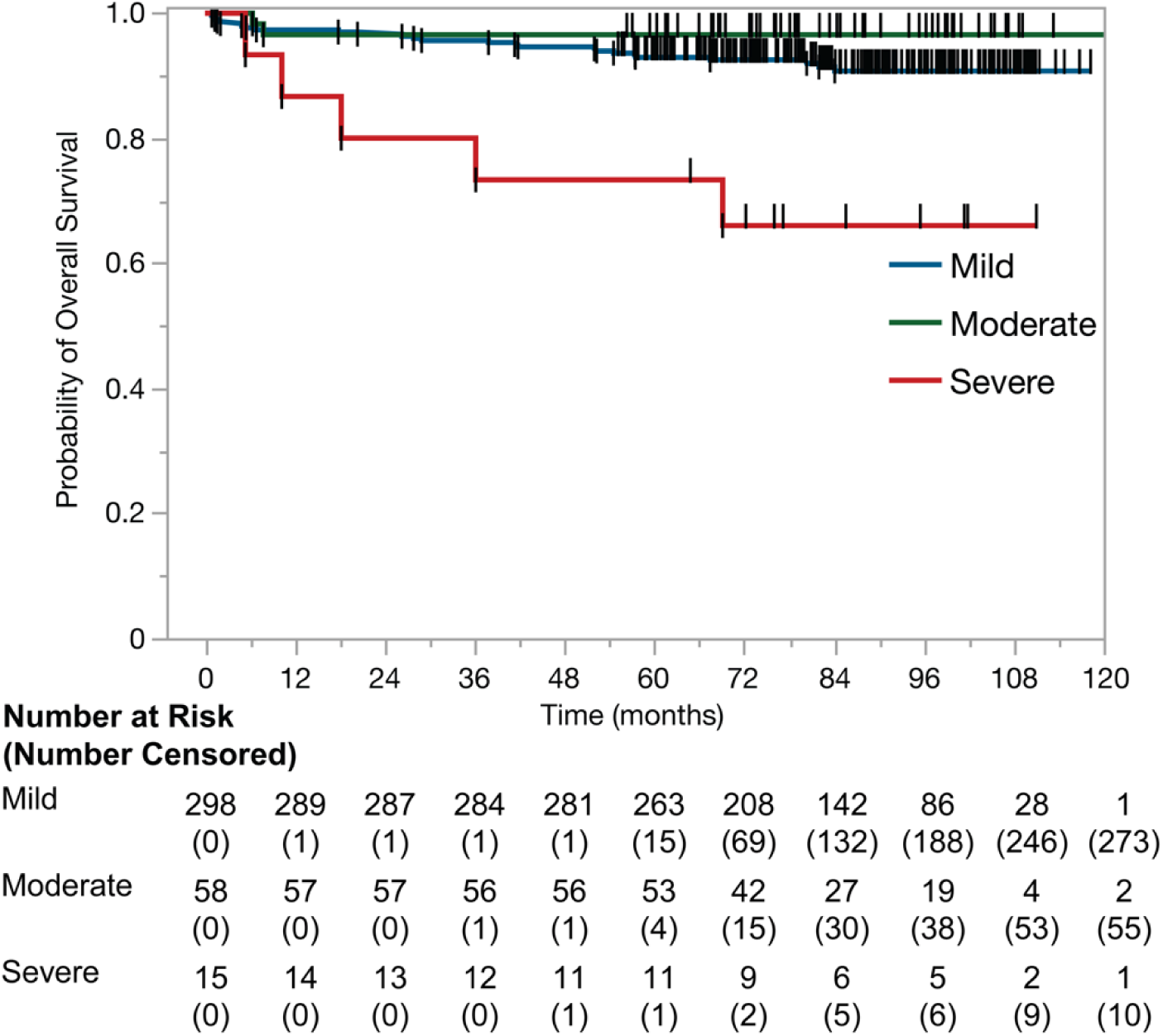
Kaplan-Meier survival curves stratified by severity of average daily sleep disturbance

### 3.4 Drowsiness

Drowsiness also increased during treatment and then improved after RT (Figure 7, S8). At baseline, most patients (88%) reported mild drowsiness, whereas 8% reported moderate, and 5% severe (mean score 1.3, SD 2.2). At the start of RT, drowsiness remained relatively low (mean score 1.4); 89% reported mild drowsiness, 8% moderate, and 4% severe. Peak drowsiness occurred at the end of RT with a mean MDASI score of 3.8 (SD 2.71). At this time, severe drowsiness was reported by 18%. After RT, drowsiness levels substantially improved. By 6 months post-RT, only 2% of patients had severe drowsiness, 9% had moderate, and 89% had mild, nearly back to baseline levels (mean score 1.3 at 6 months, 1.1 at 12 months).

**Figure 7.**
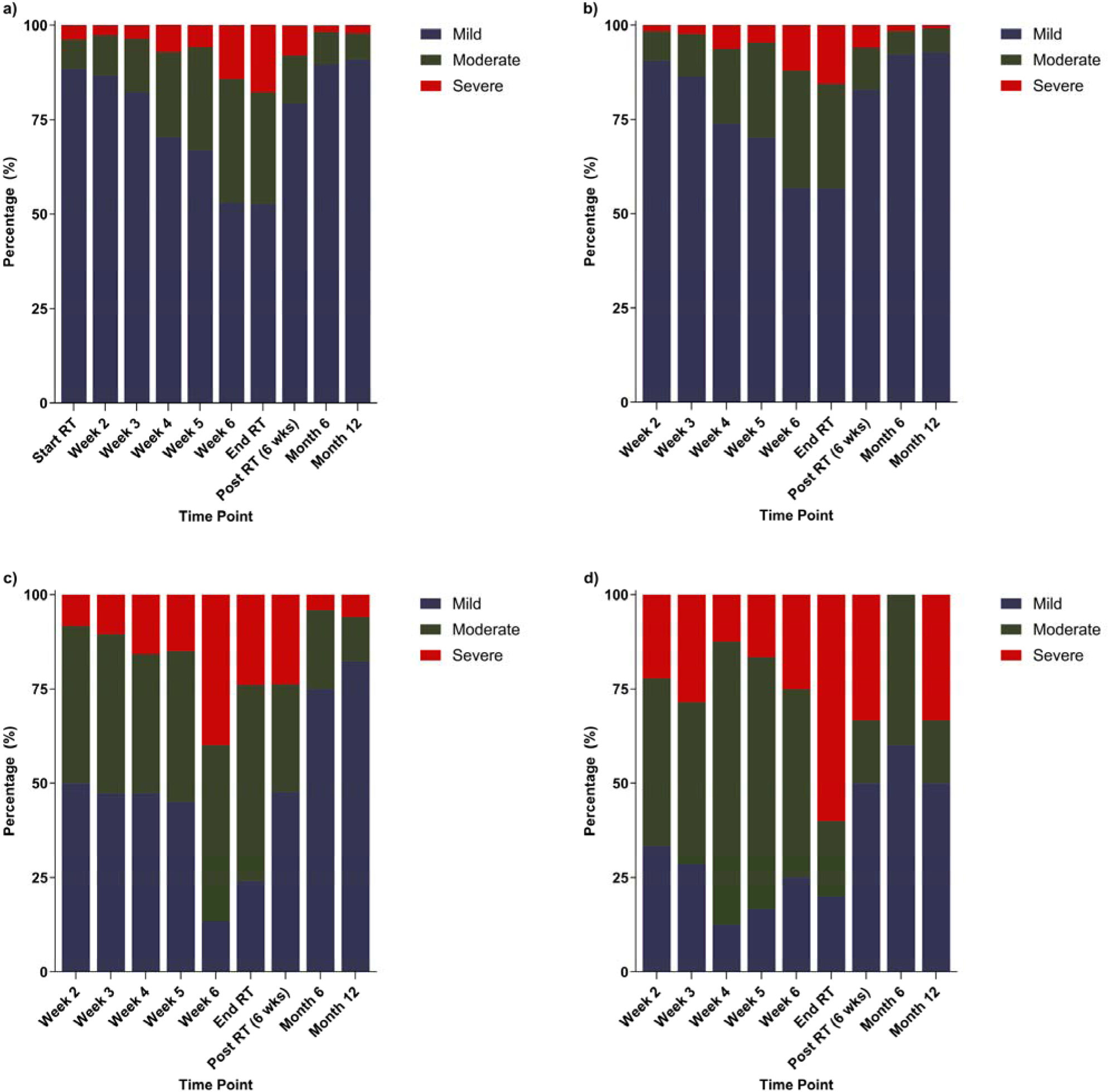
Evolution of drowsiness severity at relevant timepoints, stratified by symptom severity reported at RT start. Panels represent: (a) the entire cohort, (b) patients with mild drowsiness at RT start, (c) patients with moderate drowsiness at RT start, and (d) patients with severe drowsiness at RT start.

Patients reporting mild drowsiness at the start of treatment largely remained in the mild category across timepoints (Figure 4). Those reporting moderate and severe drowsiness experienced greater symptom fluctuation, with the greatest proportion of severe symptoms occurring at end of RT.

Severe drowsiness reported at the start of RT was also independently associated with significantly worse OS (HR: 7.76, CI: 1.45-33.47, p = 0.0193; Table 6). This finding is consistent with the observed associations between severe fatigue, sleep disturbance, and adverse survival outcomes over time. As with other symptom domains, no other variable, aside from age, was independently associated with worse OS, including severe or moderate drowsiness at baseline, end of RT, or other post-treatment timepoints. In multivariate Cox analysis of average daily drowsiness scores, severe longitudinal drowsiness burden was also significantly associated with worse survival compared with mild burden (HR 8.24, 95% CI 1.22 to 32.62, p = 0.008), while moderate burden was not significant (HR 1.39, 95% CI 0.57 to 3.03, p = 0.4349; Table 7). Aside from age, no other variables were significantly associated with OS in this model. The Kaplan-Meier curves for drowsiness severity at RT start and average daily drowsiness severity are shown in Figure 8 and Figure 9, respectively.

**Figure 8.**
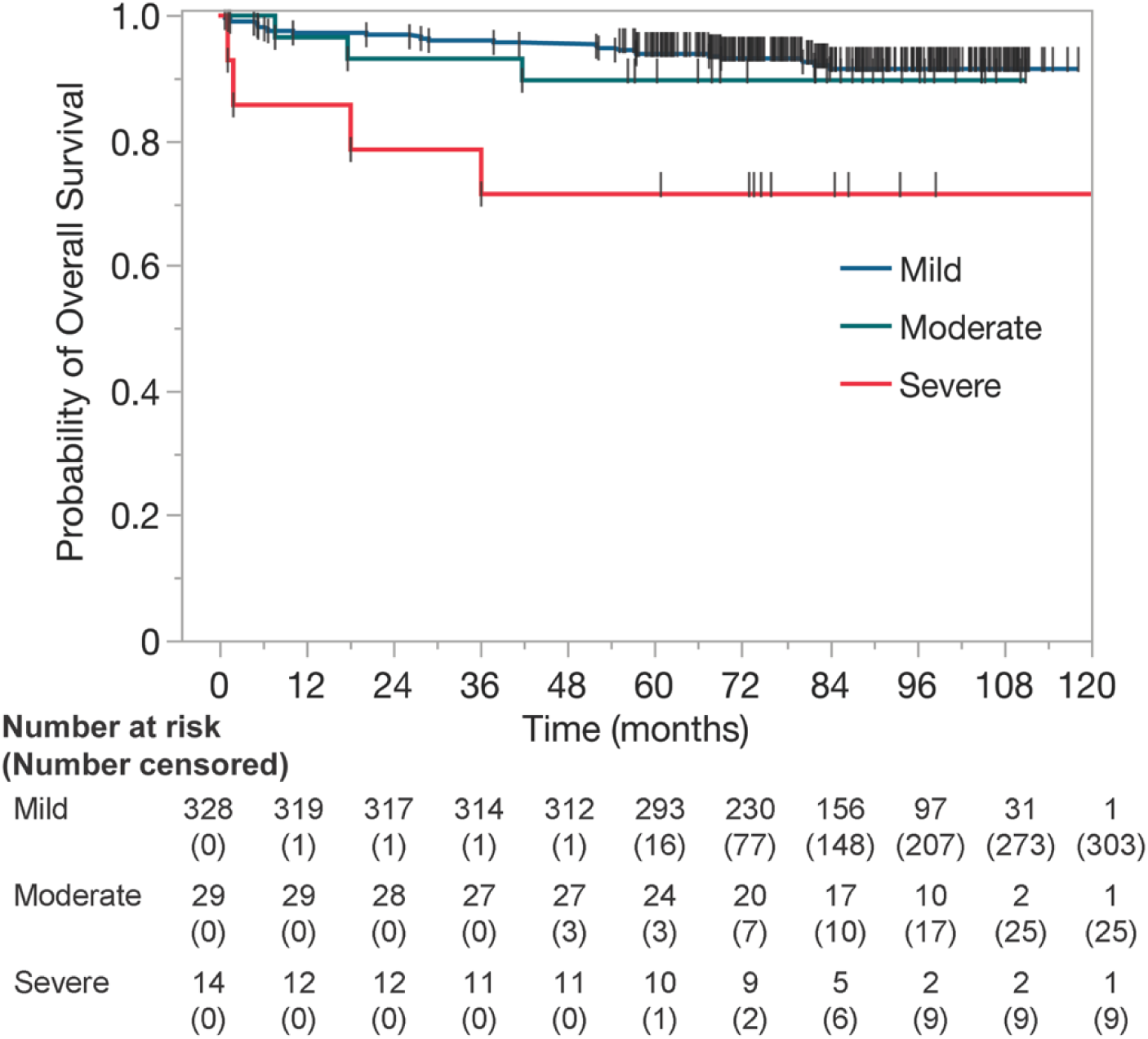
Kaplan-Meier survival curves stratified by drowsiness severity at the start of RT

**Figure 9.**
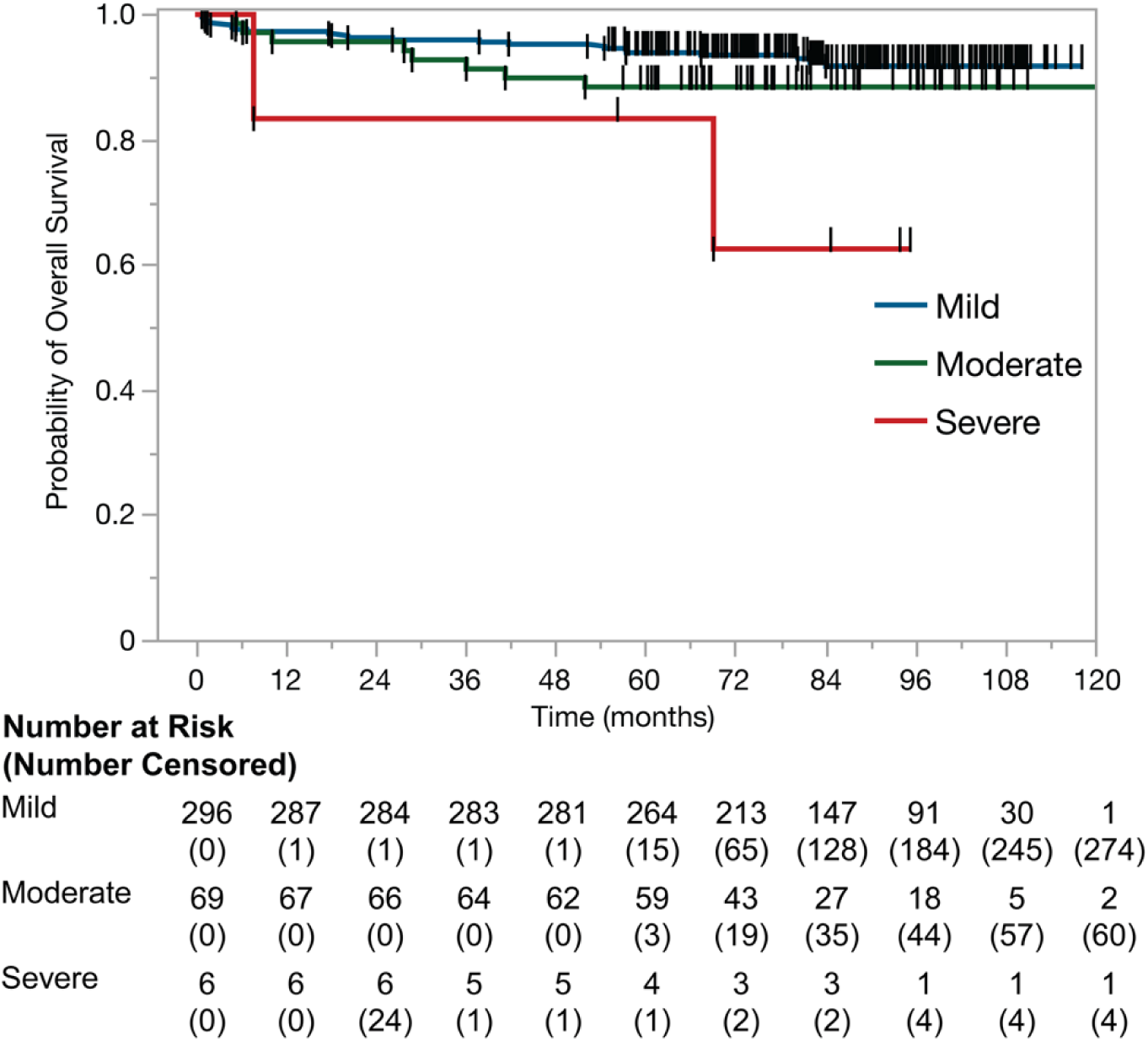
Kaplan-Meier survival curves stratified by severity of average daily drowsiness

**Table 6.** Multivariate Cox proportional hazards analysis for average daily drowsiness.

| **Variable** | **HR Estimate** | **95% CI** | **P value** |
| --- | --- | --- | --- |
| **Age** | 1.06 | 1.02-1.10 | 0.0083* |
| **Sex** |  |  |  |
| Male vs female | 1.80 | 0.50-11.73 | 0.4475 |
| **Cancer T-stage** |  |  |  |
| T1 vs T0 | 0.44 | 0.10-2.33 | 0.2994 |
| T2 vs T0 | 0.74 | 0.22-3.40 | 0.6566 |
| T3 vs T0 | 0.85 | 0.20-4.34 | 0.8342 |
| T4 vs T0 | 2.18 | 0.54-11.17 | 0.2994 |
| **Cancer n-status** |  |  |  |
| Node negative vs node positive | 0.68 | 0.12-2.34 | 0.6095 |
| **Induction chemotherapy** | 0.84 | 0.28-2.23 | 0.7308 |
| **Surgery** | 0.56 | 0.13-1.72 | 0.3704 |
| **Average daily drowsiness severity** |  |  |  |
| Moderate vs mild | 1.39 | 0.57-3.03 | 0.4349 |
| Severe vs mild | 8.24 | 1.22-32.62 | 0.0083* |

**Table 7.** Multivariate Cox proportional hazards analysis for combined average daily symptom scores.

| Variable | HR Estimate | 95% CI | P value |
| --- | --- | --- | --- |
| **Age** | 1.07 | 1.02-1.13 | 0.0047* |
| **Sex** |  |  |  |
| Male vs female | 2.15 | 0.50-16.28 | 0.3775 |
| **Cancer T-stage** |  |  |  |
| T1 vs T0 | 0.49 | 0.10-2.64 | 0.3670 |
| T2 vs T0 | 0.84 | 0.24-4.00 | 0.7998 |
| T3 vs T0 | 0.87 | 0.18-4.74 | 0.8600 |
| T4 vs T0 | 1.58 | 0.30-9.58 | 0.5958 |
| **Cancer n-status** |  |  |  |
| Node negative vs node positive | 1.30 | 0.20-4.84 | 0.7311 |
| **Induction chemotherapy** | 0.71 | 0.18-2.31 | 0.5976 |
| **Surgery** | 0.60 | 0.14-1.81 | 0.4226 |
| **Combined average daily symptoms** |  |  |  |
| Moderate vs mild | 0.88 | 0.29-2.23 | 0.7993 |
| Severe vs mild | 14.25 | 2.76-57.25 | 0.0004* |

### 3.5 Combined Average Daily Symptom Scores

In a Cox model using the combined average daily symptom scores for fatigue, sleep disturbance, and drowsiness, severe cumulative symptom burden was significantly associated with reduced OS compared with those with mild burden (HR 14.25, 95% CI 2.76 to 57.25, p = 0.0004), whereas the severe fatigue relative to moderate fatigue was not significant (HR 0.88, 95% CI 0.29 to 2.23, p = 0.7993; Table 8). Aside from age, no other variable was significantly associated with survival. Kaplan-Meier curves for combined average daily symptom scores stratified by severity (mild, moderate, severe) are shown in Figure 10.

**Figure 10.**
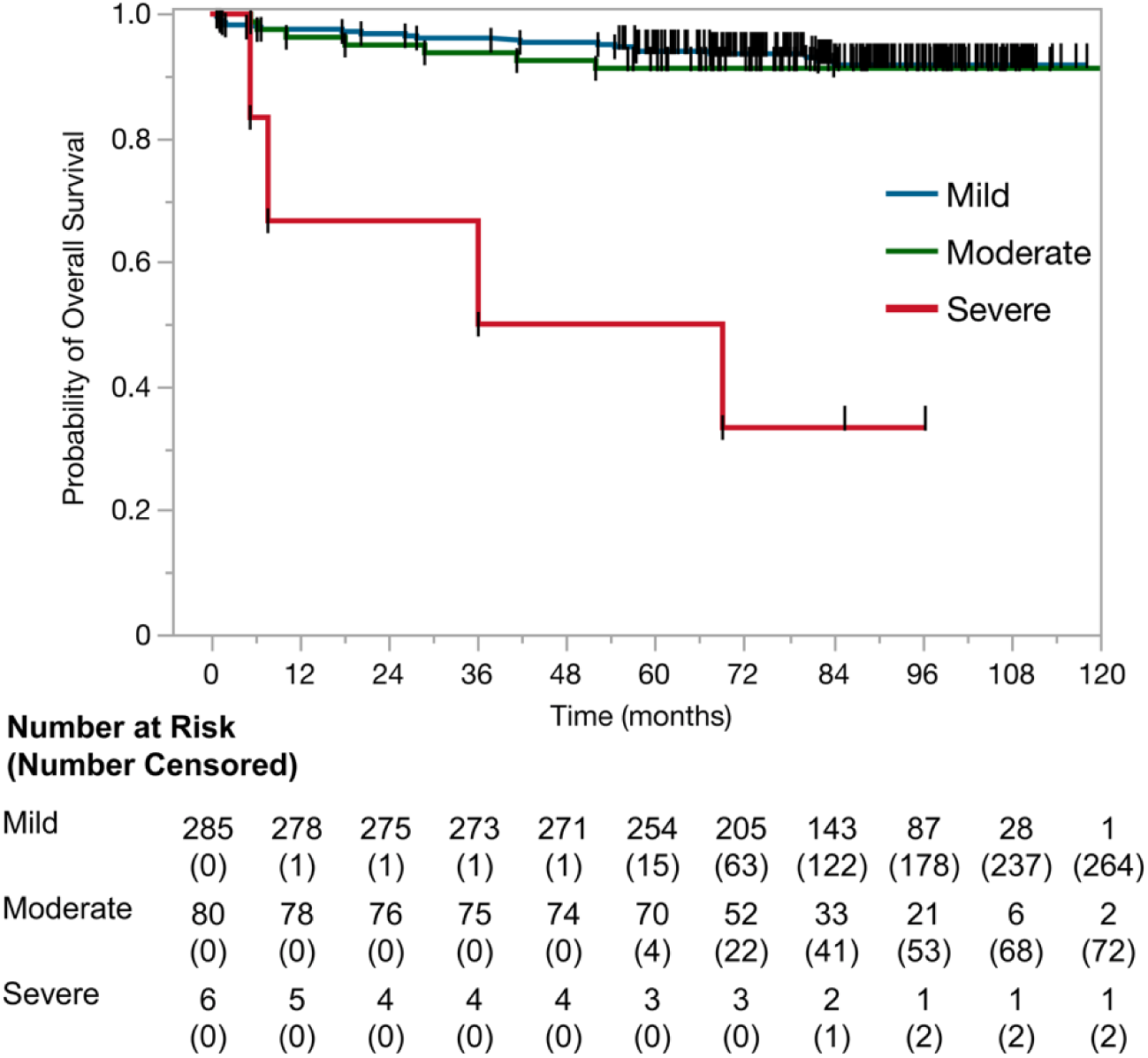
Kaplan-Meier survival curves stratified by severity of the average daily scores for all three symptoms (fatigue, sleep, drowsiness)

### 3.6 Normalized AUC Symptom Scores

Each patient’s normalized AUC score was quantified by calculating the area under the symptom trajectory curve across all relevant timepoints divided by the maximum possible area (i.e. a constant score of 10 at every time point).20 The heat map in Figure 11a shows the relationship between symptom severity reported by each patient at the start of treatment and normalized toxicity burden over the full 18 month follow up period. Toxicities (fatigue, sleep, drowsiness) on the y-axis are stratified by respective symptom severity reported at RT start on the y-axis. Each panel contains rows, with each row representing a single patient, and each row’s color corresponds to that patient’s percent of maximum AUC (ranging from 0% (blue) to 50% (red)).

**Figure 11.**
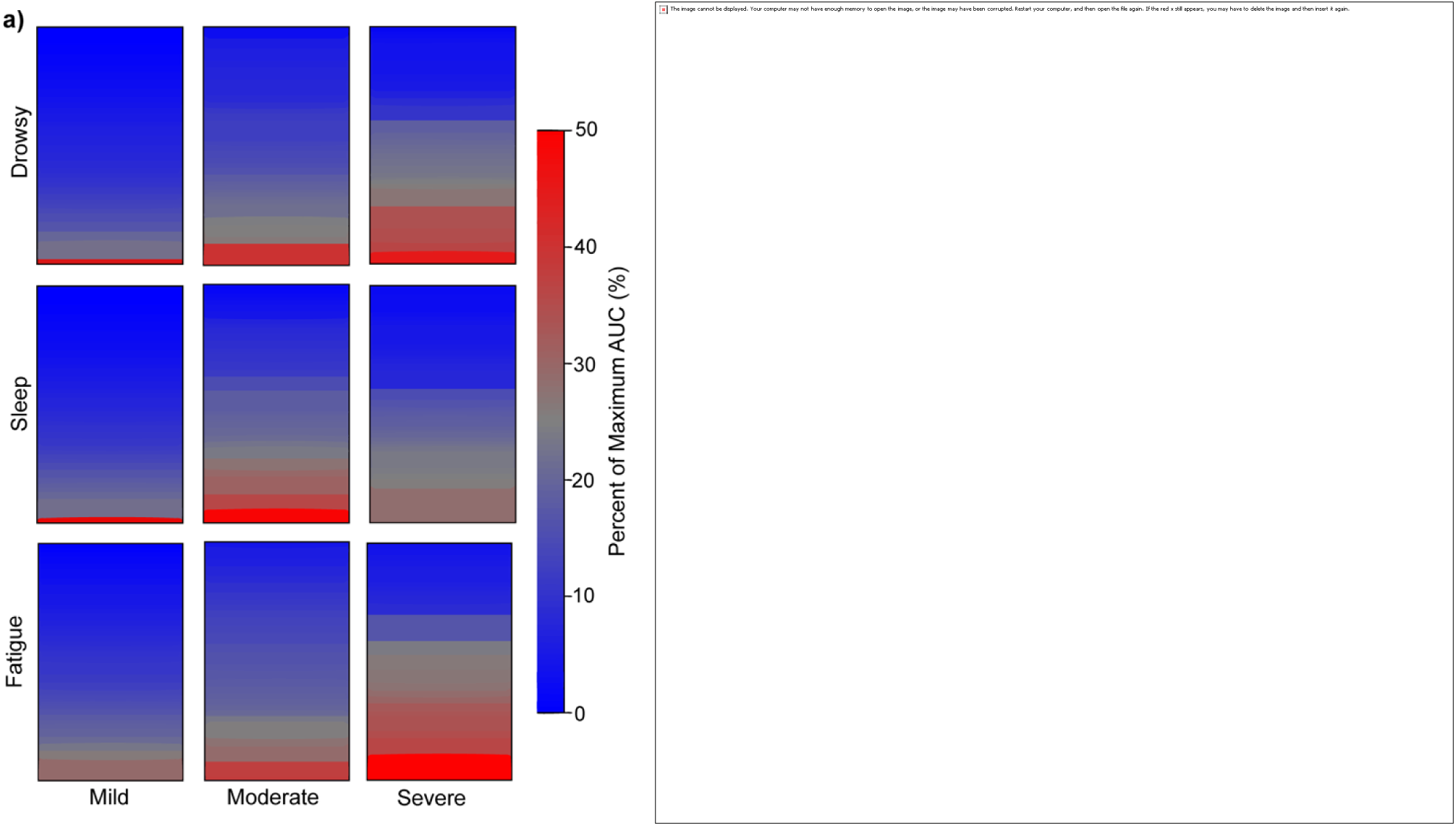
Normalized AUC scores for fatigue, sleep disturbance and drowsiness stratified by symptom severity at the start of RT

Cumulative distribution curves depicting the proportion of patients exceeding a given burden threshold for patients reporting mild (blue), moderate (grey) or severe (red) fatigue, sleep, and drowsy symptoms at RT start are shown in Figure 11b-d. The yl1laxis represents the percent of maximum symptom burden, and the xl1laxis represents the cumulative percentage of patients with an AUC at least as high as the plotted value. Figures 11e-g plot the percent of maximum AUC on the yl1laxis against successive followl1lup cutl1loff points on the xl1laxis.

Scatter plots showing comparisons of MDASI-HN symptom scores demonstrate that sleep disturbance was moderately correlated with drowsiness (r2= 0.49, Figure 12a). Sleep disturbance had a similar correlation coefficient with respect to fatigue (r2= 0.55, Figure 12b). The strongest correlation was observed between drowsiness and fatigue (r2= 0.63, Figure 12c).

**Figure 12.**
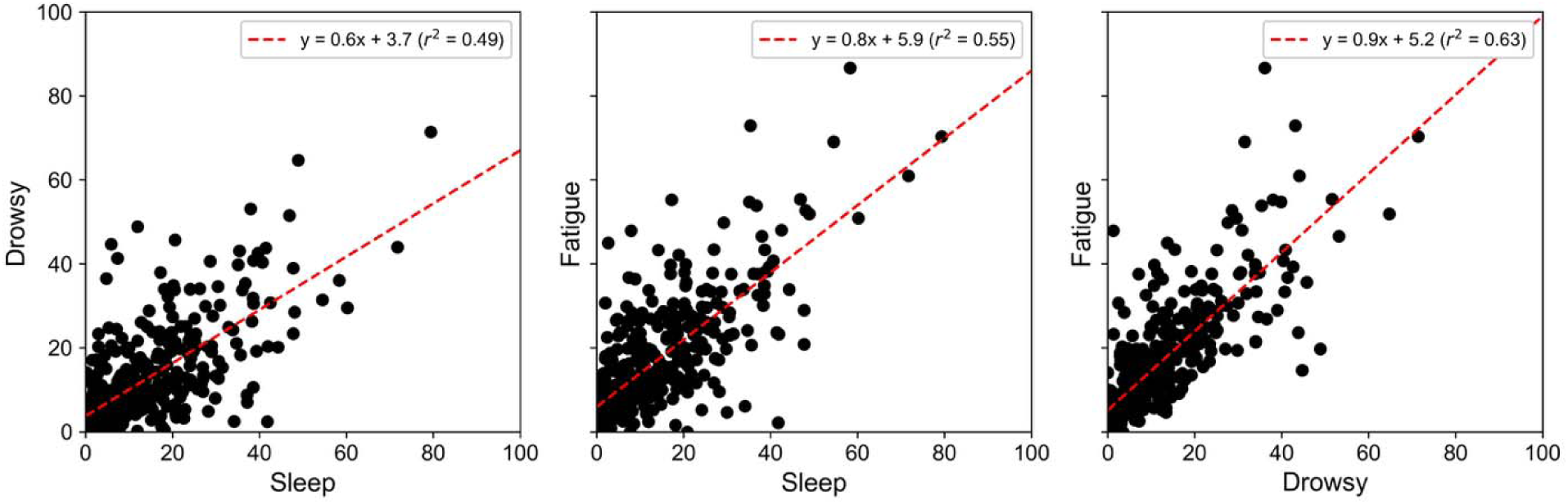
Relationship between fatigue, sleep, and drowsiness symptoms

## 4 Discussion

Our study reveals severe longitudinal sleep, fatigue, and drowsiness burdens were all independently associated with significantly worse OS at an average follow-up of 84 months in patients with OPC. Severe sleep disturbance, fatigue, and drowsiness reported at the start of RT were also independently associated with worse OS. These findings represent a significant advancement in understanding the impact of symptomatology on patient prognosis and align with our broader initiative aimed at developing targeted interventions to address symptom burden and improve outcomes in patients with OPC. To our knowledge, this is the first study to specifically examine sleep and drowsiness items in addition to fatigue in relation to survival in a cohort of patients of this type. By directly assessing these three distinct yet interconnected symptoms, we provide a comprehensive analysis of sleep-related QOL and oncologic outcomes.

In our cohort, severe sleep-related symptoms were uncommon at baseline or RT start. Other studies have shown poor sleep prior to treatment onset in patients with head and neck cancer. A multicenter study from the Netherlands by Santoso and colleagues reported poor sleep quality based on the Pittsburgh Sleep Quality Index Score (PSQI) in 44% of patients prior to start of treatment in those with head and neck cancer (n=590). Comparatively, they had a higher percentage of patients with advanced disease (stage IV disease), and their analysis showed a higher association with poor sleep in those with younger age, female sex, a passive coping style, greater oral pain, and reduced sexual interest and enjoyment. In another study evaluating sleep in patients with multiple cancers, Zhou and associates demonstrated that 59.6% of head and neck cancer patients (n=344) had a sleep disturbance prior to treatment using an electronic screening survey.^21^ In contrast, our cohort included mostly men with early-stage disease, and use of a validated survey tool longitudinally demonstrated that a substantial proportion of patients experienced clinically meaningful sleep-related disturbances by the end of RT.

Aside from T0 stage, no other variables were significantly associated with increased sleep burden in this cohort. The etiology for these sleep disturbances could range from poor sleep hygiene to an underlying sleep disorder, such as insomnia or obstructive sleep apnea (OSA). A recent meta-analysis on patients with head and neck cancer noted that insomnia affects roughly 30-45% before and during therapy and remains prevalent (around 40%) after treatment.^22^ As noted in the literature above, these patients may have had pre-existing insomnia; they could have also developed insomnia that is perpetuated by maladaptive behaviors, prolonging their insomnia symptoms well into survivorship.^22^ Use of MDASI-HN questionnaires in our cohort can help identify patients with sleep disruption at any point and subsequently provide them with tools to address their sleep issues or enable appropriate referrals to sleep specialists.

The importance of sleep health has been recognized by the American Heart Association, and it is closely and bidirectionally linked to cardiovascular health.^23^ Specifically, poor sleep health could be characterized by abnormal sleep duration, poor quality or underlying sleep disorders. Further, in patients with cancer, sleep disruption resulting in hypoxia or sleep fragmentation have also been shown to contribute to cancer progression and worse outcomes. Further study in those with head and neck cancer is needed.^24,25^

Our results suggest that severe initial symptom severity, even before RT-induced toxicity develops, can identify a vulnerable subset of patients with markedly worse survival. In our multivariate analysis, reporting a symptom as “severe” (score ≥7) at RT initiation corresponded to a 5-fold higher risk of death for sleep disturbance, a 3.5-fold higher risk for fatigue, and an approximately 8-fold higher risk for drowsiness, compared to patients with only mild or no symptoms at that time. Correspondingly, our study demonstrates that aggregate sleep related symptomatic burden is associated with worse OS, both on an individual symptom basis or in aggregate.

Our findings align with and extend upon prior research on PRO and survival. A comprehensive review by Montazeri et al.demonstrated that baseline fatigue levels can be a significant independent predictor of survival in several cancer types, such as lung, breast and gastroesophageal cancers.^26^ In patients with OPC specifically, another study reported that a 10-point increase in baseline fatigue score was associated with a 17% reduction in the likelihood of survival.^27^ Our study did not find a significant association for baseline fatigue scores in patients with OPC; however, we observed a significant association for fatigue at RT start, which could effectively serve as a “new baseline” after any initial interventions, such as surgery or induction chemotherapy, and immediately before the physiological stress of RT. This distinction could explain why baseline symptoms measured at diagnosis or pre-treatment were not prognostic in our cohort, whereas the symptoms measured on day one of RT were. While not specifically examined in this study, there is also ongoing research into possible links between sleep-related symptoms and sarcopenia, which is a known predictor of worse OS in multiple cancers.^28,29^

Drowsiness is another symptom frequently experienced by patients with OPC, though it remains underreported. Fatigue and drowsiness often overlap but are conceptually distinct. Drowsiness implies a propensity to fall asleep, whereas cancer-related fatigue is a more pervasive exhaustion not relieved by rest.^11^ In our linear regression model, prior surgical resection was significantly associated with lower drowsiness burden (coefficient = 0.21, 95% CI: 0.00 to 0.42, p = 0.049; S3), whereas age, sex, tumor site, tumor stage, nodal status, and induction chemotherapy were not significant. Prior studies have shown a pooled prevalence of 16% before treatment and 32% following curative treatment.^22^ Chronic pain induced by RT is also found in almost 70% of survivors of head and neck cancer^15^, complicating sleep/fatigue symptom profiles, as many patients may be either in acute distress or on analgesic medications. Thus, use of pain medications or limited sleep duration due to multiple factors can also lead to drowsiness. However, the direct association of drowsiness with survival outcomes has been less frequently explored.

We acknowledge several limitations. The focus of our investigation was on symptoms, so subsequent diagnostic sleep evaluation including assessment of sleep duration and underlying sleep disorders was not performed. Although OSA occurs with increased incidence in head and neck cancer,^30^ most of our cohort has sleep-related symptoms return close to baseline, so less likely had significant OSA. Additionally, it is often difficult to distinguish sleep disturbance from fatigue and drowsiness, as these interrelated symptoms overlap but may reflect distinct biological or behavioral processes, but use of the same survey tool longitudinally likely helped to discern true symptomatology. Patients lost to follow-up or missing PRO data at later timepoints might differ in outcomes, although our primary analysis focused on baseline and RT-start data where completion rates were high, and loss of data was minimal due to the use of electronic medical records.

Our data leverages a large, homogenous cohort of OPC patients with standardized treatment and long follow-up period (median 84 months), and it allows a unique view into variability of sleep-related symptoms as before, during and after treatment. Our well-defined OPC cohort limits confounding by ensuring similar tumor sites and RT fields across patients. Our use of a validated PRO instrument and established timepoints for symptom severity increases interpretability of the results. Although the biological mechanisms linking head and neck radiation to patient-reported sleep disturbances remain unclear, our findings highlight an important area for future investigation into the complex interplay between symptom burden and treatment effects.

In conclusion, we present a large, longitudinal assessment of fatigue, drowsiness, and sleep quality in a well-defined cohort of OPC patients receiving RT. Our study highlights the critical importance of assessing sleep disturbances as well as their potential physiological correlates, such as sleep apnea, hypopnea, and sarcopenia, as a part of comprehensive care.

Future efforts will focus on differentiating true sleep dysfunction from fatigue-related dysfunction by incorporating formal polysomnography and objective measures of sarcopenia and cachexia. Such approaches may clarify whether anatomical or treatment-related changes serve as drivers of these symptoms and whether they can function as prognostic indicators in OPC. In parallel, it will also be important to examine whether adverse events during RT are linked to persistently low PROs and clinical outcomes in this cohort. Early identification of high-risk patients may enable targeted interventions to improve both quality of life and survival in this population.

## Data Availability

In accordance with NOT-OD-21-013, Final NIH Policy for Data Management and Sharing, anonymized/de-identified data that support the findings of this study are openly available in an NIH-supported generalist scientific data repository (figshare) at https://doi.org/10.6084/m9.figshare.29971432.v1 no later than the time of an associated publication.

---

**Supplemental Table 1.** Multiple linear regression for average daily fatigue scores.

*Supplemental 1 Multiple linear regression for average daily fatigue scores*
| Term | Estimate | Std Error | t Ratio | Prob>\|t\| | Lower 95% | Upper 95% |
| --- | --- | --- | --- | --- | --- | --- |
| **Intercept** | 1.90 | 0.80 | 2.37 | 0.0185* | 0.32 | 3.48 |
| **Age** | 0.00 | 0.01 | 0.35 | 0.73 | -0.02 | 0.03 |
| **Sex** | 0.19 | 0.19 | 1.02 | 0.31 | -0.18 | 0.56 |
| **Tumor Site** |  |  |  |  |  |  |
| **Base of Tongue** | 0.78 | 0.36 | 2.15 | 0.0321* | 0.07 | 1.49 |
| **GPS** | -1.10 | 0.91 | -1.21 | 0.23 | -2.90 | 0.69 |
| **Nasopharynx** | -0.16 | 1.12 | -0.14 | 0.89 | -2.37 | 2.05 |
| **NOS** | -0.95 | 0.65 | -1.46 | 0.15 | -2.22 | 0.33 |
| **Pharyngeal wall** | -0.38 | 0.96 | -0.40 | 0.69 | -2.26 | 1.50 |
| **Soft palate** | 1.10 | 0.83 | 1.34 | 0.18 | -0.52 | 2.73 |
| **Cancer T-stage** |  |  |  |  |  |  |
| **T0** | 1.58 | 0.61 | 2.58 | 0.0105* | 0.37 | 2.78 |
| **T1** | -0.24 | 0.37 | -0.65 | 0.52 | -0.97 | 0.49 |
| **T2** | -0.04 | 0.35 | -0.10 | 0.92 | -0.73 | 0.65 |
| **T3** | 0.04 | 0.40 | 0.10 | 0.92 | -0.74 | 0.82 |
| **T4** | -0.31 | 0.47 | -0.65 | 0.52 | -1.24 | 0.62 |
| **Cancer n-status** |  |  |  |  |  |  |
| **Node negative** | -0.06 | 0.21 | -0.29 | 0.77 | -0.48 | 0.36 |
| **Induction Chemo** | 0.30 | 0.17 | 1.80 | 0.07 | -0.03 | 0.63 |
| **Surgery** | 0.21 | 0.13 | 1.63 | 0.10 | -0.04 | 0.46 |

**Supplemental Table 2.** Multiple linear regression for average daily sleep scores.

*Supplemental 2 Multiple linear regression for average daily sleep scores*
| **Term** | **Estimate** | **Std Error** | **t Ratio** | **Prob>\|t\|** | **Lower**<br>**95%** | **Upper**<br>**95%** |
| --- | --- | --- | --- | --- | --- | --- |
| **Intercept** | 2.43 | 0.71 | 3.41 | 0.0007* | 1.03 | 3.83 |
| **Age** | -0.01 | 0.01 | -0.55 | 0.58 | -0.02 | 0.01 |
| **Sex** | 0.24 | 0.16 | 1.55 | 0.12 | -0.07 | 0.55 |
| **Tumor Site** |  |  |  |  |  |  |
| **Base of**<br>**Tongue** | 0.38 | 0.31 | 1.22 | 0.22 | -0.23 | 0.99 |
| **GPS** | -1.07 | 0.76 | -1.40 | 0.16 | -2.57 | 0.43 |
| **Nasopharynx** | -0.04 | 1.05 | -0.04 | 0.97 | -2.11 | 2.03 |
| **NOS** | -0.94 | 0.57 | -1.65 | 0.10 | -2.07 | 0.18 |
| **Pharyngeal**<br>**wall** | 0.68 | 0.72 | 0.95 | 0.34 | -0.73 | 2.10 |
| **Soft palate** | 0.69 | 0.78 | 0.89 | 0.37 | -0.84 | 2.22 |
| **Cancer T-stage** |  |  |  |  |  |  |
| **T0** | 1.11 | 0.56 | 1.99 | 0.0472* | 0.01 | 2.21 |
| **T1** | -0.41 | 0.34 | -1.21 | 0.23 | -1.09 | 0.26 |
| **T2** | -0.24 | 0.33 | -0.72 | 0.47 | -0.88 | 0.41 |
| **T3** | -0.22 | 0.36 | -0.60 | 0.55 | -0.92 | 0.49 |
| **T4** | -0.22 | 0.40 | -0.55 | 0.58 | -1.01 | 0.57 |
| **Cancer n-**<br>**status** |  |  |  |  |  |  |
| **Node negative** | 0.02 | 0.18 | 0.14 | 0.89 | -0.33 | 0.37 |
| **Induction**<br>**Chemo** | 0.16 | 0.15 | 1.11 | 0.27 | -0.13 | 0.45 |
| **Surgery** | 0.20 | 0.11 | 1.77 | 0.08 | -0.02 | 0.43 |

**Supplemental Table 3.** Multiple linear regression for average daily drowsiness scores.

*Supplemental 3 Multiple linear regression for average daily drowsiness scores*
| Term | Estimate | Std Error | t Ratio | Prob>\|t\| | Lower 95% | Upper 95% |
| --- | --- | --- | --- | --- | --- | --- |
| **Intercept** | 1.45 | 0.67 | 2.17 | 0.0304* | 0.14 | 2.76 |
| **Age** | 0.00 | 0.01 | 0.45 | 0.66 | -0.01 | 0.02 |
| **Sex** | 0.23 | 0.15 | 1.60 | 0.11 | -0.05 | 0.52 |
| **Tumor Site** |  |  |  |  |  |  |
| **Base of Tongue** | 0.32 | 0.29 | 1.08 | 0.28 | -0.26 | 0.89 |
| **GPS** | -0.99 | 0.71 | -1.39 | 0.17 | -2.39 | 0.41 |
| **Nasopharynx** | -0.57 | 0.99 | -0.58 | 0.56 | -2.51 | 1.37 |
| **NOS** | -0.43 | 0.54 | -0.81 | 0.42 | -1.49 | 0.62 |
| **Pharyngeal wall** | 0.68 | 0.67 | 1.00 | 0.32 | -0.65 | 2.00 |
| **Soft palate** | 0.75 | 0.73 | 1.03 | 0.30 | -0.68 | 2.19 |
| **Cancer T-stage** |  |  |  |  |  |  |
| **T0** | 0.74 | 0.52 | 1.42 | 0.16 | -0.29 | 1.77 |
| **T1** | -0.09 | 0.32 | -0.27 | 0.79 | -0.72 | 0.54 |
| **T2** | 0.06 | 0.31 | 0.20 | 0.84 | -0.54 | 0.66 |
| **T3** | 0.17 | 0.34 | 0.51 | 0.61 | -0.49 | 0.84 |
| **T4** | -0.02 | 0.37 | -0.06 | 0.95 | -0.76 | 0.71 |
| **Cancer n-status** |  |  |  |  |  |  |
| **Node negative** | -0.02 | 0.17 | -0.14 | 0.89 | -0.35 | 0.30 |
| **Induction Chemo** | 0.14 | 0.14 | 1.04 | 0.30 | -0.13 | 0.41 |
| **Surgery** | 0.21 | 0.11 | 1.97 | 0.0494* | 0.00 | 0.42 |

**Supplemental Table 4.**
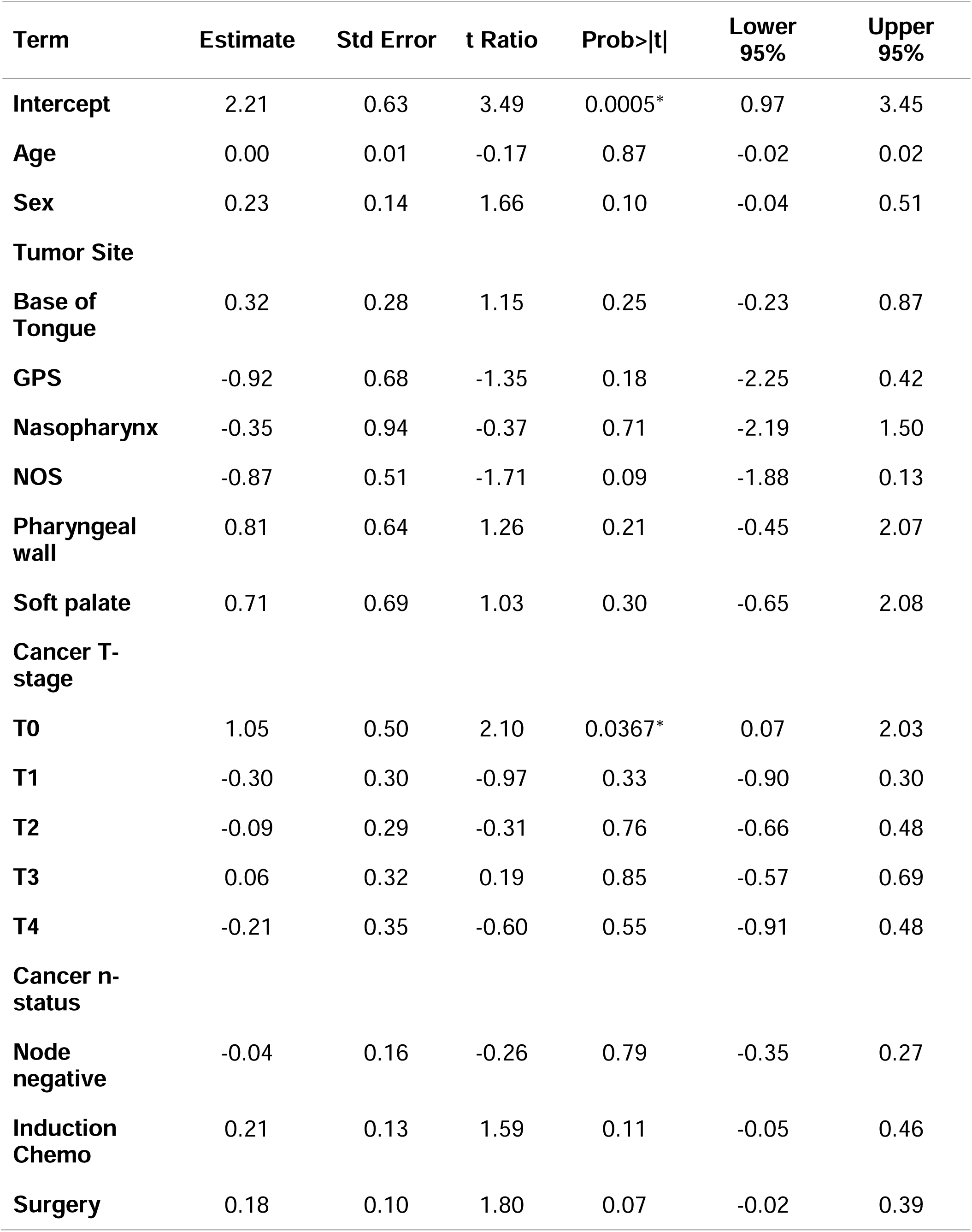
Multiple linear regression for combined average daily symptom scores.

**Supplemental Table 5.** Mean MDASI-HN Scores across all symptom categories.

*Supplemental 5 Mean MDASI-HN Scores across all symptom categories*
|  | **Baseline** | **Start RT** | **End RT** | **Post RT (6 wks)** | **6 Months** | **12 Months** |
| --- | --- | --- | --- | --- | --- | --- |
|  | Mean score (SD) | Mean Score (SD) | Mean Score (SD) | Mean Score (SD) | Mean Score (SD) | Mean Score (SD) |
|  | n = 312 | n = 372 | n = 292 | n = 260 | n = 285 | n = 235 |
| **Fatigue** | 1.8 (2.25) | 1.9 (2.11) | 4.5 (2.57) | 3.0 (2.39) | 1.9 (2.06) | 1.6 (1.90) |
| **Sleep** | 1.6 (2.32) | 1.7 (2.11) | 3.5 (2.65) | 2.4 (2.41) | 1.5 (1.95) | 1.4 (1.97) |
| **Drowsy** | 1.3 (2.16) | 1.4 (1.92) | 3.8 (2.71) | 2.0 (2.30) | 1.3 (1.63) | 1.1 (1.61) |

**Supplemental Table 6.**
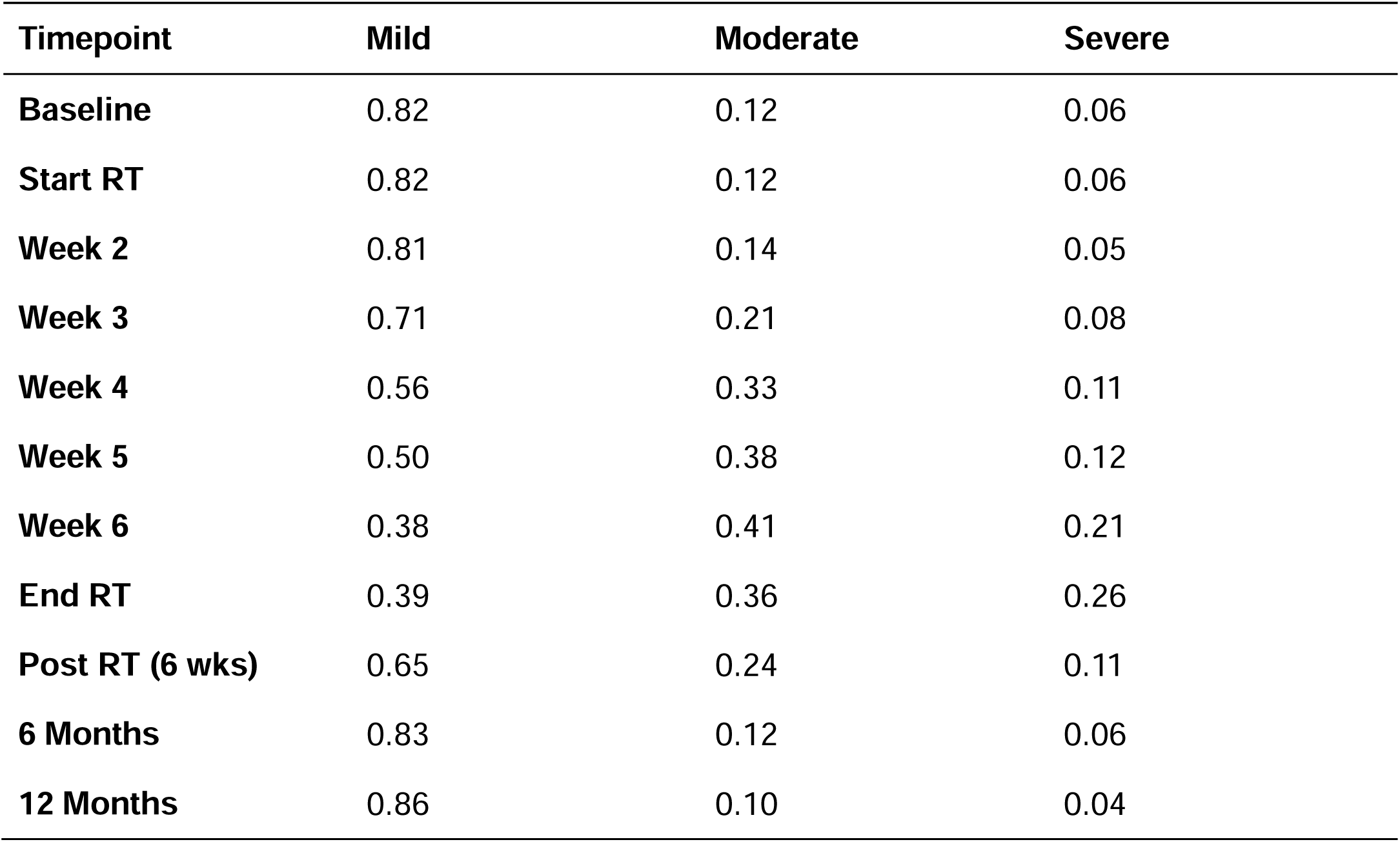
Severity Distribution of Fatigue Symptomatology.

**Supplemental Table 7.**
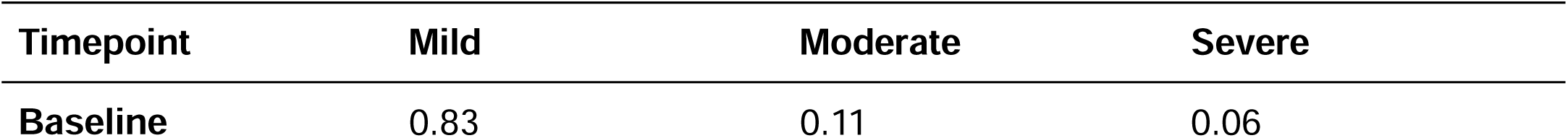
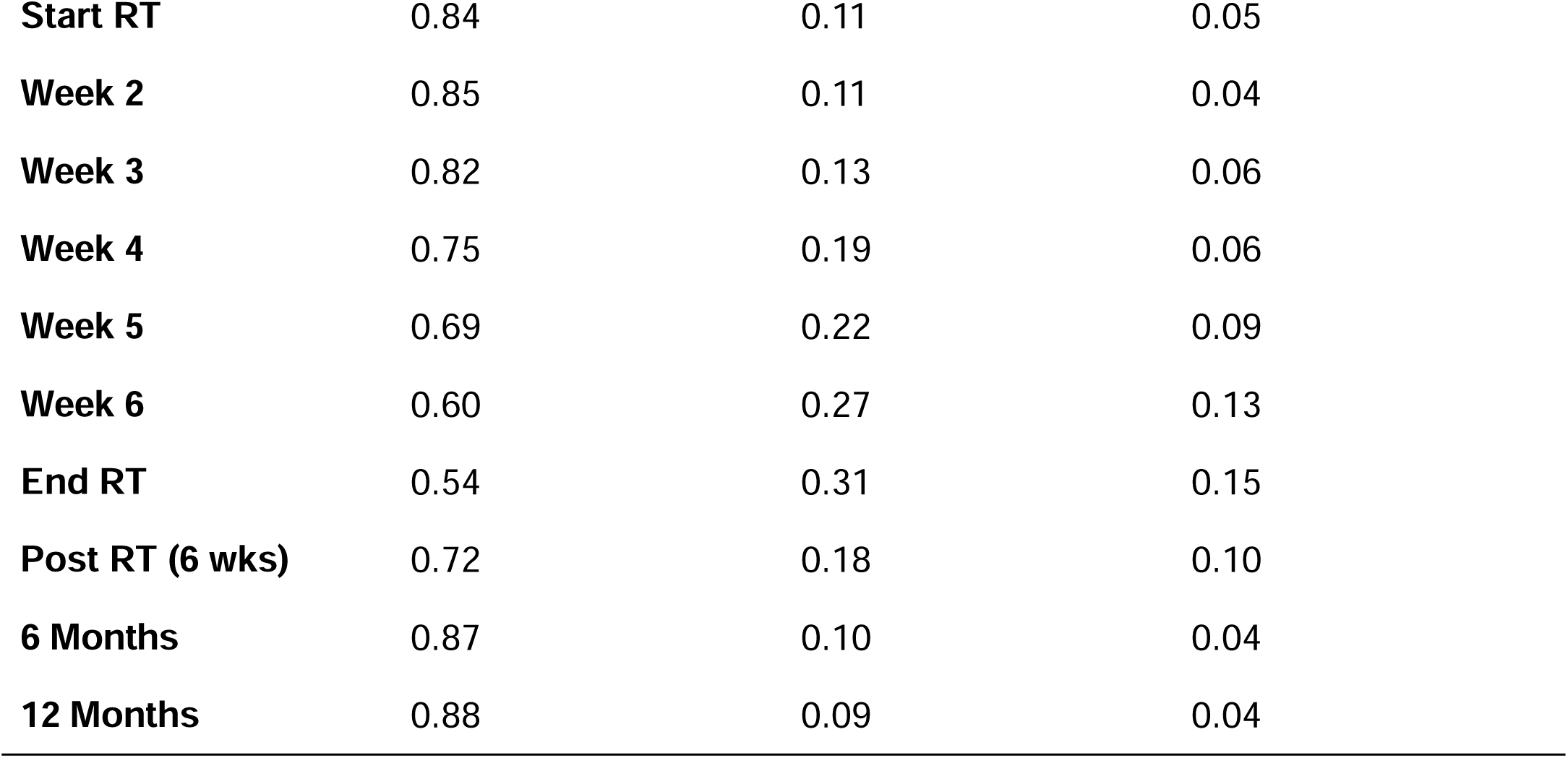
Severity Distribution of Sleep Symptomatology.

**Supplemental Table 8.** Severity Distribution of Drowsiness Symptomatology.

*Supplemental 8 Severity Distribution of Drowsiness Symptomatology*
| **Timepoint** | **Mild** | **Moderate** | **Severe** |
| --- | --- | --- | --- |
| **Baseline** | 0.88 | 0.08 | 0.05 |
| **Start RT** | 0.89 | 0.08 | 0.04 |
| **Week 2** | 0.87 | 0.11 | 0.03 |
| **Week 3** | 0.82 | 0.14 | 0.04 |
| **Week 4** | 0.71 | 0.23 | 0.07 |
| **Week 5** | 0.68 | 0.27 | 0.06 |
| **Week 6** | 0.53 | 0.33 | 0.14 |
| **End RT** | 0.53 | 0.30 | 0.18 |
| **Post RT (6 wks)** | 0.79 | 0.13 | 0.08 |
| **6 Months** | 0.89 | 0.09 | 0.02 |
| **12 Months** | 0.91 | 0.07 | 0.02 |

**Supplemental Table 9.** Multivariate Cox proportional hazards analysis for fatigue (baseline, end RT)

*Supplemental 9 Multivariate Cox proportional hazards analysis for fatigue (baseline, end RT)*
| **Variable** | **HR Estimate** | **95% CI** | **P value** |
| --- | --- | --- | --- |
| **Age** | 1.12 | 1.05-1.21 | 0.0005 |
| **Sex** |  |  |  |
| **Male vs female** | 8.63E-12 | -- | 0.2806 |
| **Cancer T-stage** |  |  |  |
| **T1 vs T0** | 0.73 | 0.06-18.45 | 0.8171 |
| **T2 vs T0** | 0.91 | 0.12-19.78 | 0.9350 |
| **T3 vs T0** | 1.15 | 0.12-26.67 | 0.9125 |
| **T4 vs T0** | 4.86 | 0.35-144.4 | 0.2511 |
| **Cancer n-status** |  |  |  |
| **Node negative vs node positive** | 3.33E-12 | -- | 0.0334* |
| **Induction chemotherapy** | 0.86 | 0.14-4.95 | 0.8722 |
| **Surgery** | 0.40 | 0.02-2.58 | 0.3752 |
| **Baseline Fatigue Severity** |  |  |  |
| **Moderate vs mild** | 0.21 | 0.01-1.40 | 0.1187 |
| **Severe vs mild** | 3.00E-12 | -- | 0.2091 |
| **Start RT Fatigue severity** |  |  |  |
| **Moderate vs mild** | 4.14 | 1.17-19.62 | 0.0261* |
| **Severe vs mild** | 1.49 | 0.19-9.41 | 0.6720 |

**Supplemental Table 10.** Multivariate Cox proportional hazards analysis for sleep (baseline, end RT)

*Supplemental 10 Multivariate Cox proportional hazards analysis for sleep (baseline, end RT)*
| **Variable** | **HR Estimate** | **95% CI** | **P value** |
| --- | --- | --- | --- |
| **Age** | 1.11 | 1.04-1.20 | 0.0021* |
| **Sex** |  |  |  |
| **Male vs female** | 0.9.39E-12 | -- | 0.2471 |
| **Cancer t-stage** |  |  |  |
| **t1 vs t0** | 1.06 | 0.09-24.84 | 0.9662 |
| **t2 vs t0** | 1.10 | 0.15-22.77 | 0.9362 |
| **t3 vs t0** | 1.93 | 0.22-41.80 | 0.5721 |
| **t4 vs t0** | 8.33 | 0.91-188.9 | 0.0608 |
| **Cancer n-status** |  |  |  |
| **Node negative vs node positive** | 3.55E-12 | -- | 0.0417* |
| **Induction chemotherapy** | 0.80 | 0.13-3.56 | 0.7873 |
| **Surgery** | 0.30 | 0.02-1.72 | 0.2052 |
| **Baseline sleep severity** |  |  |  |
| **Moderate vs mild** | 0.35 | 0.02-2.36 | 0.3132 |
| **Severe vs mild** | 6.90E-12 | -- | 0.1942 |
| **End RT sleep severity** |  |  |  |
| **Moderate vs mild** | 1.65 | 0.23-7.46 | 0.5715 |
| **Severe vs mild** | 1.02 | 0.22-3.67 | 0.9823 |

**Supplemental Table 11.** Multivariate Cox proportional hazards analysis for drowsiness (baseline, end RT)

*Supplemental 11 Multivariate Cox proportional hazards analysis for drowsiness (baseline, end RT)*
| **Variable** | **HR Estimate** | **95% CI** | **P value** |
| --- | --- | --- | --- |
| **Age** | 1.11 | 1.04-1.20 | 0.0029* |
| **Sex** |  |  |  |
| **Male vs female** | 7.9e-12 | -- | 0.2467 |
| **Cancer t-stage** |  |  |  |
| **t1 vs t0** | 0.91 | 0.07-22.17 | 0.9410 |
| **t2 vs t0** | 1.01 | 0.15-20.71 | 0.9917 |
| **t3 vs t0** | 1.53 | 0.17-22.66 | 0.7243 |
| **t4 vs t0** | 4.85 | 0.47-116.7 | 0.1942 |
| **Cancer n-status** |  |  |  |
| **Node negative vs node positive** | 4.7e-12 | -- | 0.0757 |
| **Induction chemotherapy** | 0.94 | 0.15-4.80 | 0.9438 |
| **Surgery** | 0.31 | 0.02-1.69 | 0.2038 |
| **Baseline drowsiness severity** |  |  |  |
| **Moderate vs mild** | 2.42E-12 | -- | 0.1584 |
| **Severe vs mild** | 3.97E-12 | -- | 0.2681 |
| **End RT drowsiness severity** |  |  |  |
| **Moderate vs mild** | 1.53 | 0.22-7.36 | 0.6288 |
| **Severe vs mild** | 2.19 | 0.69-7.57 | 0.1808 |

